# Excess mortality in England and Scotland in 2022: The long shadow of austerity and the return to an unacceptable pre-pandemic baseline

**DOI:** 10.1101/2025.03.03.25323230

**Authors:** Daniel R. R. Bradford, Denise Brown, Gerard McCartney, Margaret Douglas, Ruth Dundas, David Walsh

## Abstract

**Background:** There are concerns that mortality remains elevated after peaks COVID-19. This study examined whether mortality rates in England and Scotland in 2022 were excessive compared to rates predicted by austerity-era (2012–2019) and pre-austerity (2001–2010) trends.

**Methods:** A linear time trend analysis was conducted using mortality data from 2001–2022. The outcomes were observed and expected age- and sex-standardised mortality rates (ASMRs; standardised to the 2013 European Standard Population). Expected ASMRs in 2022 were calculated independently based on austerity-era and pre-austerity trends. Excess deaths were estimated by comparing observed and expected ASMRs.

**Results:** In 2022, ASMRs were higher than predicted by austerity-era trends but substantially higher than pre-austerity trends. Relative excesses in England for females were 4.4% (4.0–4.8) and 38.2% (95% CI: 37.7–38.7), respectively; for males, excesses were 7.2% (6.8–7.6) and 57.0% (56.4–57.6). Relative excesses in Scotland for females were 3.4% (2.2–4.5) and 26.6% (25.2–28.0); for males, excesses were 2.6% (1.5–3.8) and 45.2% (43.6–46.9). COVID-19 accounted for 5.3–6.5% of deaths in 2022 and explained much of the excess relative to austerity-era trends. ASMRs in the most deprived areas were 1.68–1.94 times higher than in the least deprived.

**Conclusion:** Mortality was higher than predicted by both austerity-era and pre-austerity trends. Deaths attributable to COVID-19 explain a substantial proportion of the excess based on austerity-era trends. However, the 879,430 excess deaths relative to pre-austerity trends, even after excluding direct COVID-19 deaths, highlights the devastating impacts of austerity on public health.

**KEY MESSAGES:** *What is already known on this topic?:* Mortality in England and Scotland remained elevated following the peaks of 2020 and 2021 caused by COVID-19, indicating a further worsening of the unprecedented stalling of mortality rates observed from around 2012.

*What this study adds:* While COVID-19 explained much of the 2022 excess mortality relative to projections based on austerity-era trends, far greater excesses emerged when compared to projections based on the consistent decline in mortality observed prior to austerity.

*How this study might affect research, practice or policy:* These findings highlight the profound, long-term harm of austerity, particularly in deprived areas, with an estimated 879,430 excess deaths between 2013 and 2022. The study strengthens calls for urgent policy action to reverse austerity’s effects and reduce health inequalities.

## INTRODUCTION

There are concerns that UK mortality rates remained elevated after the COVID-19 peaks in 2020 and 2021 compared to the preceding years.[1–4] Before considering potential causes of continuing increased mortality, it is essential to determine whether an excess exists. Whether a rate is deemed excessive depends on the definition of the expected counterfactual rate. In defining the counterfactual, both temporal trends in mortality before the pandemic and changes in the population’s age and sex structure must be considered. Prior studies have not always done so. For example, the Organisation for Economic Co-operation and Development (OECD) and the Office for National Statistics (ONS) used mean crude death counts from the five years before the pandemic as a baseline for estimating excess deaths.[4,5] The recently updated ONS methodology relies on only the most recent five years of data, and therefore cannot account for the major shift in mortality trends that began around 2012 in the UK.[6–10] The purpose of this study was to compare mortality in England and Scotland in 2022 with expected rates extrapolated from longer-term pre-pandemic trends in age- and sex-standardised mortality rates (ASMRs).

Any examination of recent mortality trends must consider the changes observed in the UK from around 2012 onwards.[7–11] Around this time, national mortality rates stopped declining—reversing over a century of improvement except during the World Wars and the 1918 influenza pandemic.[10,12,13] These altered trends have been seen across nearly all age groups and causes of death.[14,15] However, the impact has not been uniform: mortality rates in the most socioeconomically disadvantaged areas have increased, and inequalities between the most and least disadvantaged areas have widened.[7,10] There is now an overwhelming body of evidence attributing these unprecedented changes in mortality to the imposition of ‘austerity’ by the UK Government from 2010 onwards.[16–21] Austerity has been implemented through enormous public spending cuts, particularly affecting poorer populations via reductions in social security and the defunding of vital public services. Consequently, using trends in ASMRs in the years immediately prior to the pandemic to predict expected counterfactual ASMRs in 2022 (as is the case in ONS publications) is based on a faulty assumption i.e., that trends in period just before the pandemic were ‘normal’ and were simply a continuation of the previous trajectory. This assumption obscures the true scale of excess mortality observed since austerity was introduced. This study therefore estimated expected counterfactual ASMRs in 2022 predicted by trends observed both before and after the imposition of austerity.

Several hypotheses have been proposed to explain persistently high mortality following the COVID-19 peaks in high-income countries, as summarised in a recent scoping review.[22] One hypothesis is that the excesses are caused by direct COVID-19 deaths. This study therefore also considered the contribution of direct COVID-19 deaths in England and Scotland in 2022.

The research questions addressed were:

1. How did observed ASMRs in England and Scotland in 2022 compare to expected ASMRs predicted by a continuation of austerity-era trends (2012-2019)?
2. How did observed ASMRs in England and Scotland in 2022 compare to expected ASMRs predicted by a continuation of pre-austerity trends (2001-2010)?
3. Were differences in observed and expected ASMRs based on the two prediction periods consistent across subgroups of age, sex, and area deprivation?
4. What proportion of deaths in England and Scotland in 2022 were attributable to COVID-19?

## METHODS

This study followed REporting of studies Conducted using Observational Routinely-collected Data (RECORD) guidelines (see supplemental material for checklist).[23]

### Data

Mortality data for England and Scotland for 2001-2022 were obtained from the Office for National Statistics and National Records of Scotland.[24] Scottish data included cause of death, while English data provided only counts of deaths mentioning COVID-19 without details on other causes.[25] In this study, a COVID-19 death was defined for both nations as any death where COVID-19 was mentioned, not necessarily as the underlying cause.

Population data for England and Scotland for 2001–2022 were obtained from the Office for National Statistics and National Records of Scotland.[24] For most years, data were disaggregated by five-year age group, sex, and deprivation level. However, for 2021–2022 in England and 2022 in Scotland, population data were only available by age and sex, with no breakdown by deprivation level. To estimate counts for each age–sex–deprivation group, we assumed that the relative distribution across the five deprivation levels within each age–sex group remained unchanged from the most recent published data (2021 for England and 2021 for Scotland) and applied these proportions to the available age–sex data.

Deprivation was measured using national indices: the Index of Multiple Deprivation (IMD) for England and the Scottish Index of Multiple Deprivation (SIMD). As both are periodically updated, mortality data were linked to the chronologically-nearest available version (see Tables A1a and A1b). Although derived separately using slightly different indicators and spatial scales, the two measures have been shown to be comparable.[26] Standardised mortality rates are included in both the English and Scottish overall IMD, but previous work has shown the effect of this circularity to be negligible.[27,28] Deprivation was categorised into five groups from most to least deprived.

### Analysis

ASMRs and 95% confidence intervals were calculated using the ‘PHEindicatormethods’ package v2.0.2 in R v4.2.0, standardised to the 2013 European Standard Population (ESP2013). The upper age group comprised individuals aged 85 or older. In addition to all-age rates, ASMRs were computed for subgroups—premature mortality (0–74y), older adults (>74y), older working age (45–64y), and younger working age (20–44y). ASMRs were calculated nationally and across five deprivation levels, rate ratios were defined relative to the least deprived group.

#### Counterfactual estimation

Expected ASMRs were calculated from two baseline counterfactuals based on extended pre-pandemic trends in the pre-austerity (2001–2010) and austerity (2012–2019) periods, rather than relying on the most recent five years of data as in ONS reports. Data were plotted to check for linearity, and linear regression models were extrapolated to 2022 with year as the independent variable and ASMRs as the dependent variable. Independent models were fitted for each combination of nation, sex, age, and area deprivation to generate expected ASMRs. Confidence intervals were not calculated for expected ASMRs as the number of datapoints used in each model did not meet normality assumptions, though trends are clearly linear (see, for example, Figure A3). The start year of 2001 was chosen to provide a similar number of data years for both periods and because mortality data disaggregated by deprivation level were available from that year. The terminal year of 2010 was selected to coincide with the imposition of austerity, and 2012 was chosen based on inflection points in trends observed in 2011–2013.[7,8] A sensitivity analysis was conducted using start years of 2011, 2012, and 2013 for the austerity baseline, while 2019 was chosen to omit peak COVID-19 mortality.

#### Excess mortality

Excess mortality was defined as an observed ASMR exceeding the expected ASMR. Relative excess mortality, expressed as a percentage, equals 100*((observed ASMR – expected ASMR)/expected ASMR). Confidence intervals were based on those calculated for observed ASMRs. Expected death counts were computed by multiplying ASMRs by the relevant population and adjusting for age structure differences relative to the ESP2013. The correction factor was determined by dividing observed deaths by the product of the observed ASMR and the relevant population (see Appendix for details). This factor was calculated for each combination of nation, sex, five-year age group, area deprivation, and year. Excess deaths were estimated by subtracting expected from observed counts.

### Patient and public involvement

There was no patient or public involvement in this study, and no ethical approval was required.

## RESULTS

In 2022, relative excess ASMRs were 5.6% in England and 2.9% in Scotland compared to predictions from austerity-era trends (2012–2019), and 44.5% and 33.1% respectively compared to pre-austerity trends (2001–2010). Figure 1 shows ASMRs from 2001–2022, summarising 21,431,282 deaths in England and 2,475,589 in Scotland. Table 1 presents the related observed and expected ASMRs for 2022. Across all nations and age subgroups, males had higher ASMRs. Excesses were observed in most age subgroups except younger working age adults in Scotland (see Figures A1a–A1d in the Appendix). Males in the older working (45–64y) and older adult (>74y) groups experienced the highest relative excess mortality compared to both baseline trends.

**Figure 1.**
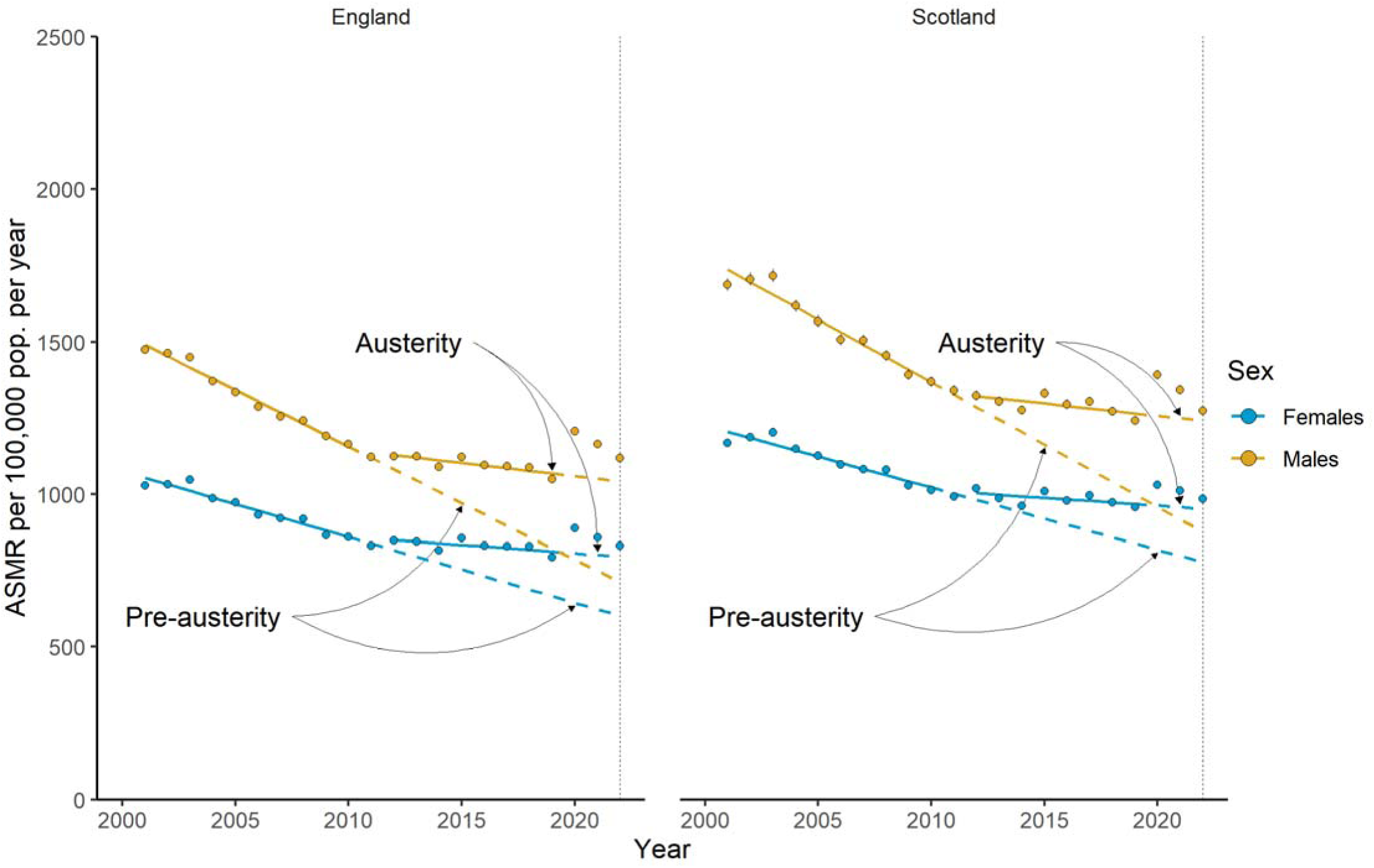
Age-standardised all-cause mortality rates from 2001–2022 in England and Scotland. Points are observed rates. Confidence intervals are included for observed rates but are so narrow as to be obscured by the points in most cases. Solid lines are trends fitted through 2001–2010 (pre-austerity) and 2012–2019 (austerity). Dashed lines are trends from each of the fitted period extrapolated to 2022.

**Table 1.** Age-standardised all-cause mortality rates per 100,000 population per year by nation, sex, and age subgroup in 2022.

| Nation | Sex | Age group | Obs. rate<br>(95% CI) | Exp. rate, 12-19 | Excess %, 12-19<br>(95%CI) | Exp. rate 01-10 | Excess %, 01-10<br>(95% CI) |
| --- | --- | --- | --- | --- | --- | --- | --- |
| England | Persons | All | 961<br>(959, 964) | 911 | 5.6<br>(5.3, 5.9) | 665 | 44.5<br>(44.1, 44.9) |
| England | Females | All | 831<br>(828, 835) | 796 | 4.4<br>(4.0, 4.8) | 602 | 38.2<br>(37.7, 38.7) |
| England | Males | All | 1118<br>(1114, 1122) | 1042 | 7.2<br>(6.8, 7.6) | 712 | 57<br>(56.4, 57.6) |
| England | Females | 0-74y | 269<br>(267, 271) | 251 | 7.1<br>(6.3, 7.9) | 183 | 47<br>(45.9, 48.1) |
| England | Males | 0-74y | 420<br>(417, 422) | 386 | 8.8<br>(8.2, 9.5) | 251 | 67.4<br>(66.3, 68.4) |
| England | Females | >74y | 2131<br>(2118, 2144) | 1999 | 6.6<br>(6.0, 7.2) | 1432 | 48.8<br>(48.0, 49.7) |
| England | Males | >74y | 3082<br>(3066, 3099) | 2867 | 7.5<br>(6.9, 8.1) | 1736 | 77.6<br>(76.6, 78.5) |
| England | Females | 45-64y | 344<br>(340, 348) | 325 | 6<br>(4.7, 7.2) | 260 | 32.5<br>(30.9, 34.1) |
| England | Males | 45-64y | 537<br>(532, 542) | 500 | 7.4<br>(6.3, 8.5) | 360 | 49.2<br>(47.7, 50.7) |
| England | Females | 25-44y | 56<br>(55, 58) | 55 | 3.2<br>(0.4, 6.0) | 46 | 23.6<br>(20.3, 27.0) |
| England | Males | 25-44y | 103<br>(100, 105) | 98 | 4.6<br>(2.4, 6.7) | 86 | 19.9<br>(17.5, 22.4) |
| Scotland | Persons | All | 1116<br>(1107, 1125) | 1084 | 2.9<br>(2.1, 3.7) | 839 | 33.1<br>(32.0, 34.1) |
| Scotland | Females | All | 985<br>(974, 996) | 953 | 3.4<br>(2.2, 4.5) | 778 | 26.6<br>(25.2, 28.0) |
| Scotland | Males | All | 1273<br>(1259, 1288) | 1240 | 2.6<br>(1.5, 3.8) | 877 | 45.2<br>(43.6, 46.9) |
| Scotland | Females | 0-74y | 347<br>(340, 354) | 337 | 3.1<br>(1.0, 5.2) | 264 | 31.4<br>(28.8, 34.1) |
| Scotland | Males | 0-74y | 514<br>(505, 523) | 503 | 2.2<br>(0.4, 4.0) | 318 | 61.7<br>(58.9, 64.5) |
| Scotland | Females | >74y | 2628<br>(2584, 2672) | 2510 | 4.7<br>(2.9, 6.4) | 2115 | 24.2<br>(22.1, 26.3) |
| Scotland | Males | >74y | 3648<br>(3592, 3705) | 3496 | 4.4<br>(2.7, 6.0) | 2440 | 49.5<br>(47.2, 51.9) |
| Scotland | Females | 45-64y | 446<br>(431, 460) | 439 | 1.5<br>(-1.8, 4.9) | 332 | 34.4<br>(30.0, 38.8) |
| Scotland | Males | 45-64y | 676 | 665 | 1.6 | 390 | 73.4 |
|  |  |  | (658, 695) |  | (-1.2, 4.5) |  | **(68.6, 78.2)** |
| Scotland | Females | 25-44y | 82 | 97 | **-15.6** | 82 | -0.6 |
|  |  |  | (76, 88) |  | **(-21.9, -9.3)** |  | (-8.0, 6.9) |
| Scotland | Males | 25-44y | 147 | 181 | **-18.6** | 149 | -1.3 |
|  |  |  | (139, 155) |  | **(-23.2, -13.9)** |  | (-6.9, 4.4) |

Notes: *Obs. rate* are observed ASMRs. *Exp. rate, 12-19* and *Exp. rate, 01-10* are the expected ASMRs if linear trends from the period 2012-2019 and 2001-2010, respectively, had continued to 2022 (see dashed lines in Figure 1). *Excess percentages* use expected rates as denominators. Bold excess values indicate results where the 95% CI of relative excess excludes zero.

### Inequalities

ASMRs increased with area deprivation in both nations and sexes. ASMRs for the most and least disadvantaged fifths from 2001–2022 are shown in Figure 2 and Tables 2a and 2b (see Figure A2 and Tables A2a and A2b for all five deprivation groups). Based on austerity-era trends, the least deprived areas had slightly larger absolute excesses than the most deprived. However, excesses based on pre-austerity trends were much greater and disproportionately affected more deprived areas. In many cases, *relative* excesses were higher in the least deprived areas — a likely artefact of more adverse prior trends in the most deprived areas predicting higher expected ASMRs. In England in 2022, ASMRs in the most deprived areas were 1.68 times higher than in the least deprived for females (up from 1.53 in 2010); the equivalent rate ratios were 1.75 (up from 1.65) for males in England, 1.94 (up from 1.80) for females in Scotland, and 1.94 (up from 1.85) for males in Scotland.

**Figure 2.**
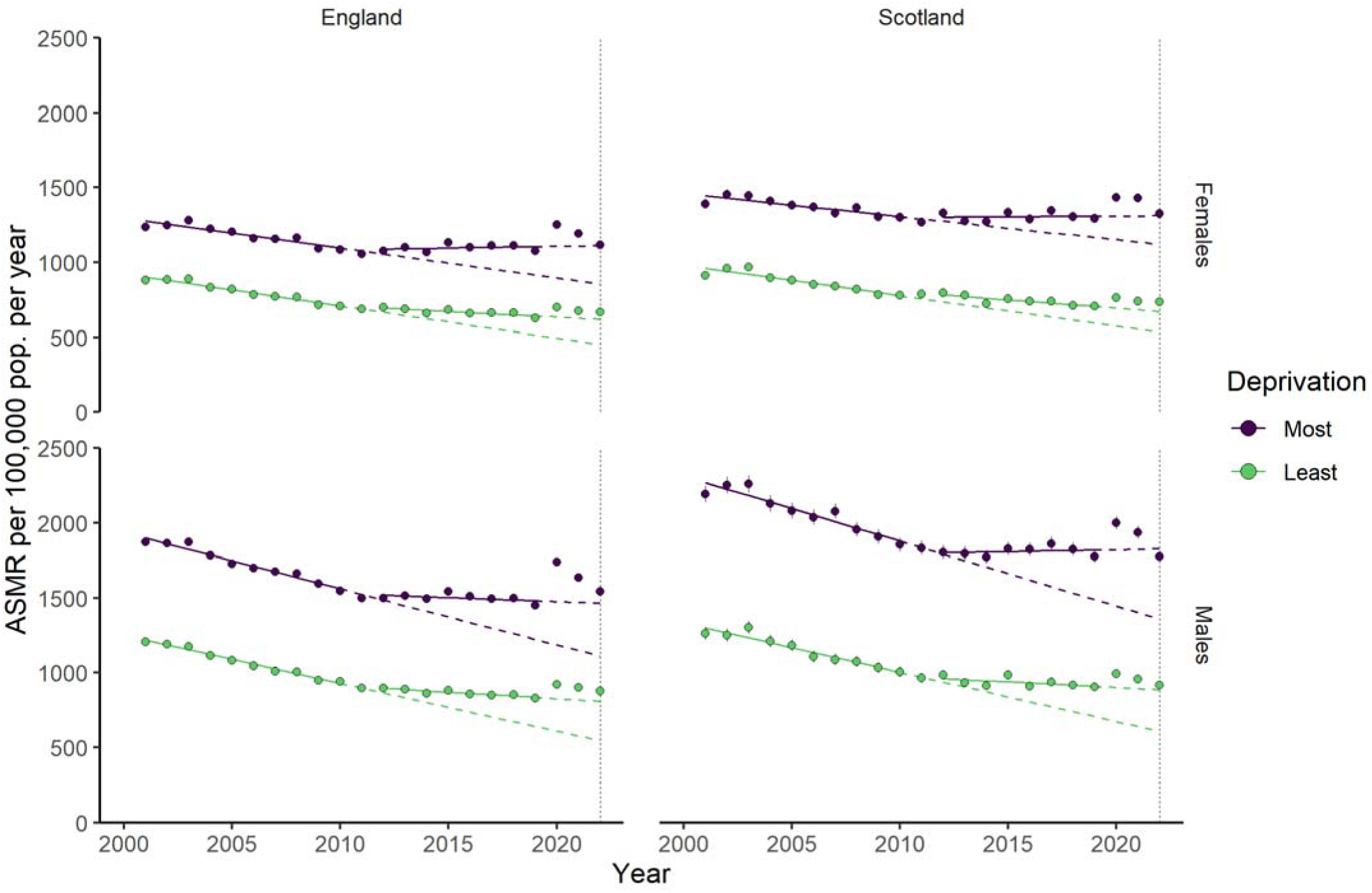
Age-standardised all-cause mortality rates from 2001–2022 in the most and least deprived fifths of the population. Points are observed rates. Confidence intervals are included for observed rates but are so narrow as to be obscured by the points in most cases. Solid lines are trends fitted through 2001–2010 (pre-austerity) and 2012– 2019 (austerity). Dashed lines are trends extrapolated to 2022.

**Table 2a.**
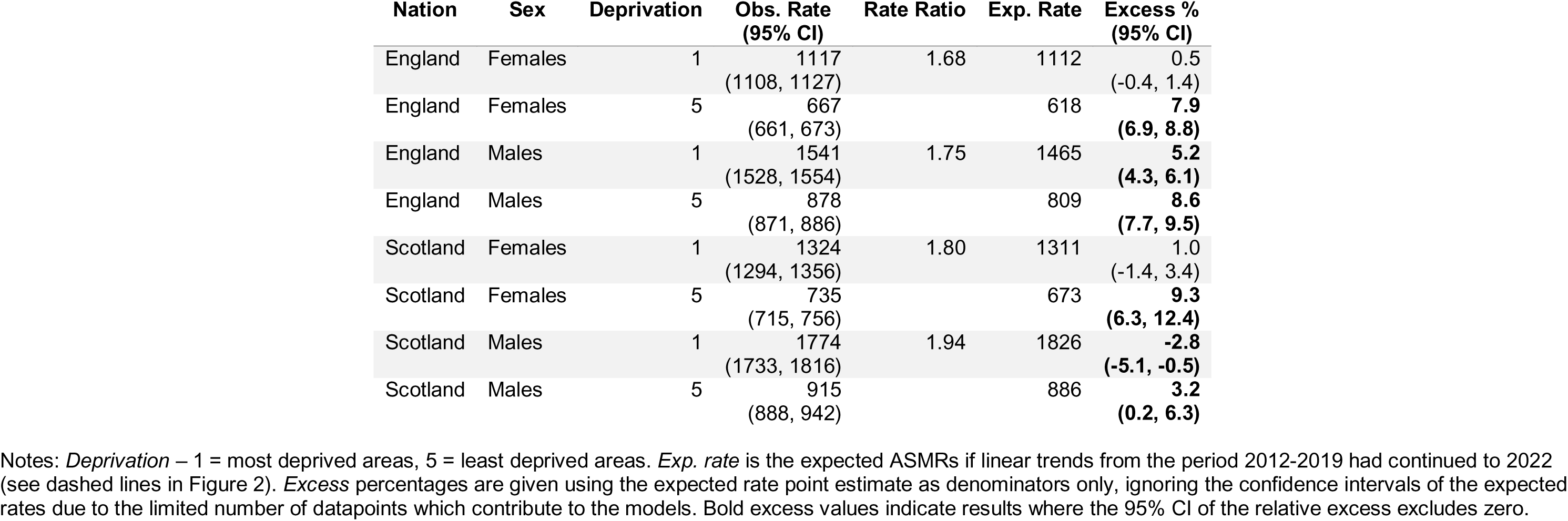
Age-standardised all-cause mortality rates per 100,000 population per year, rate ratios, and relative excess in ASMRs in 2022 in the most and least deprived areas, assuming continuation of austerity-era (2012-2019) trends.

| Nation | Sex | Deprivation | Obs. Rate<br>(95% CI) | Rate Ratio | Exp. Rate | Excess %<br>(95% CI) |
| --- | --- | --- | --- | --- | --- | --- |
| England | Females | 1 | 1117<br>(1108, 1127) | 1.68 | 1112 | 0.5<br>(-0.4, 1.4) |
| England | Females | 5 | 667<br>(661, 673) |  | 618 | **7.9**<br>**(6.9, 8.8)** |
| England | Males | 1 | 1541<br>(1528, 1554) | 1.75 | 1465 | **5.2**<br>**(4.3, 6.1)** |
| England | Males | 5 | 878<br>(871, 886) |  | 809 | **8.6**<br>**(7.7, 9.5)** |
| Scotland | Females | 1 | 1324<br>(1294, 1356) | 1.80 | 1311 | 1.0<br>(-1.4, 3.4) |
| Scotland | Females | 5 | 735<br>(715, 756) |  | 673 | **9.3**<br>**(6.3, 12.4)** |
| Scotland | Males | 1 | 1774<br>(1733, 1816) | 1.94 | 1826 | **-2.8**<br>**(-5.1, -0.5)** |
| Scotland | Males | 5 | 915<br>(888, 942) |  | 886 | **3.2**<br>**(0.2, 6.3)** |

Notes: *Deprivation* – 1 = most deprived areas, 5 = least deprived areas. *Exp. rate* is the expected ASMRs if linear trends from the period 2012-2019 had continued to 2022 (see dashed lines in Figure 2). *Excess percentages* are given using the expected rate point estimate as denominators only, ignoring the confidence intervals of the expected rates due to the limited number of datapoints which contribute to the models. Bold excess values indicate results where the 95% CI of the relative excess excludes zero.

**Table 2b.**
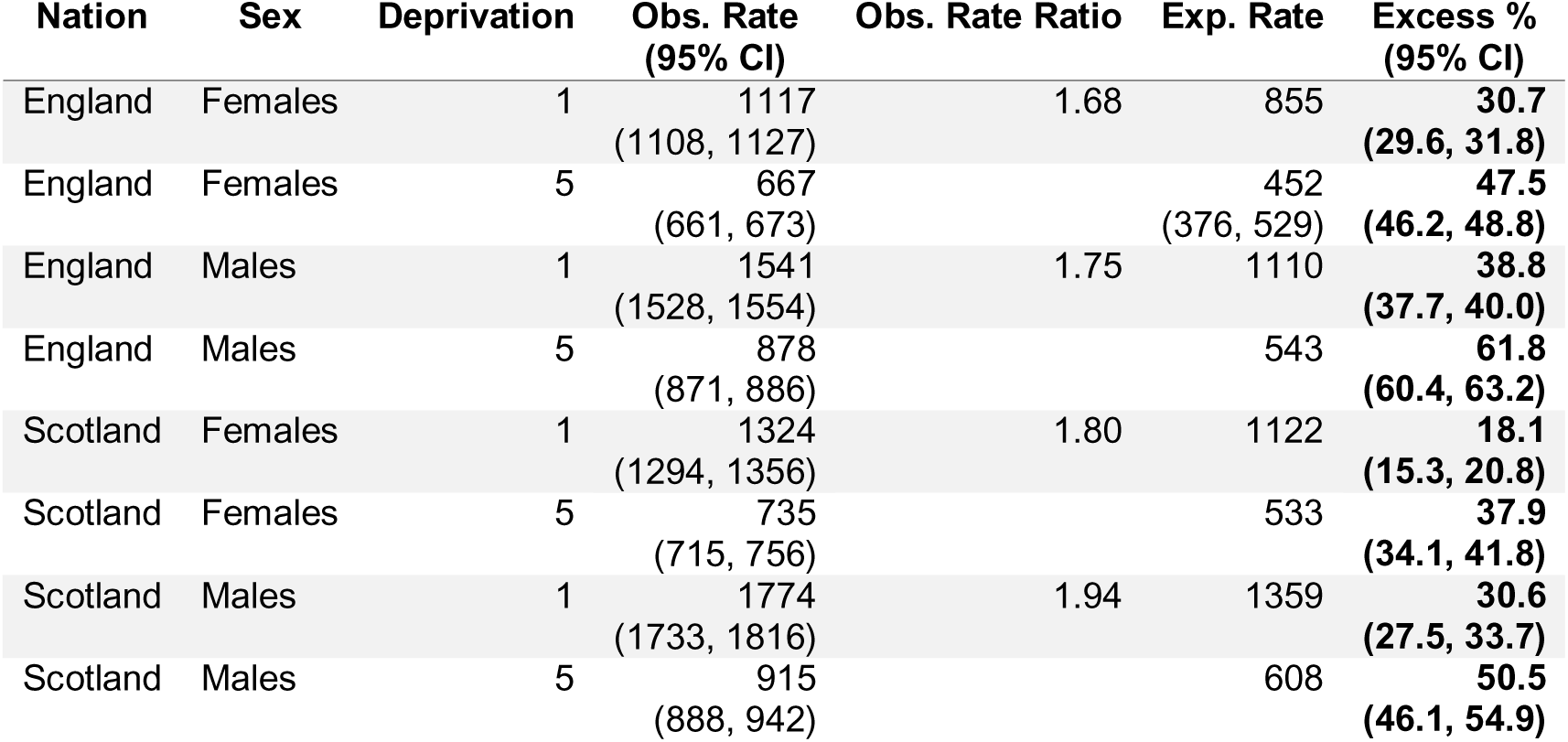
Age-standardised all-cause mortality rates per 100,000 population per year, rate ratios, and relative excess in ASMRs in 2022 in the most and least deprived areas, assuming continuation of pre-austerity (2001-2010) trends.

### The impact of excluding direct COVID-19 deaths

COVID-19 was recorded on 5.7% of death certificates in England and 6.2% in Scotland in 2022. Excluding these deaths, relative excess ASMR in England dropped from 5.6% to −0.5% and in Scotland from 2.9% to −3.5% based on austerity-era trends. These values as presented in Table 3. Across age subgroups in England, the largest remaining excess after excluding COVID-19 deaths was in premature mortality — 2.5% for females and 4.5% for males — while all Scottish age subgroups showed mortality deficits.

**Table 3.**
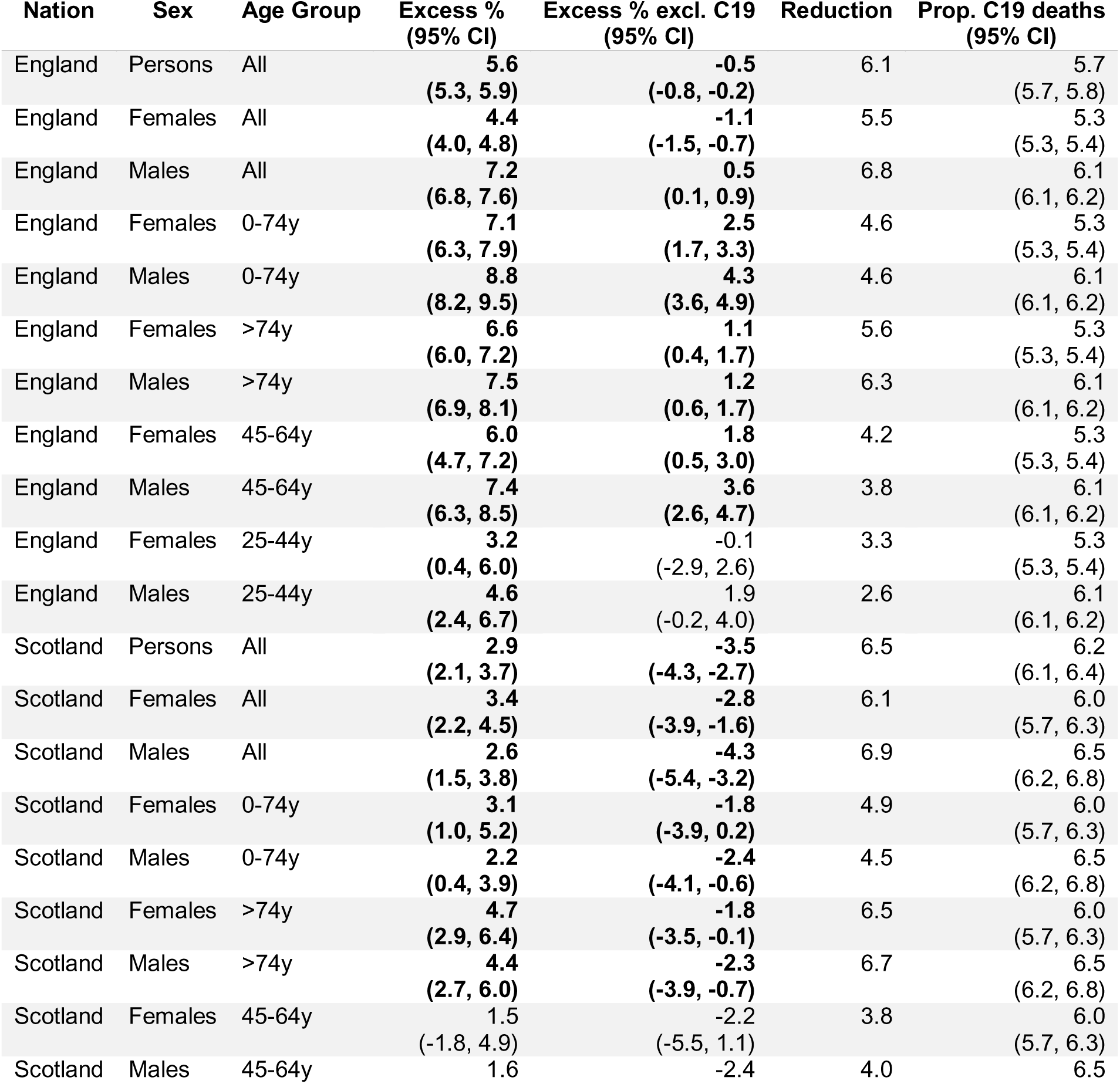
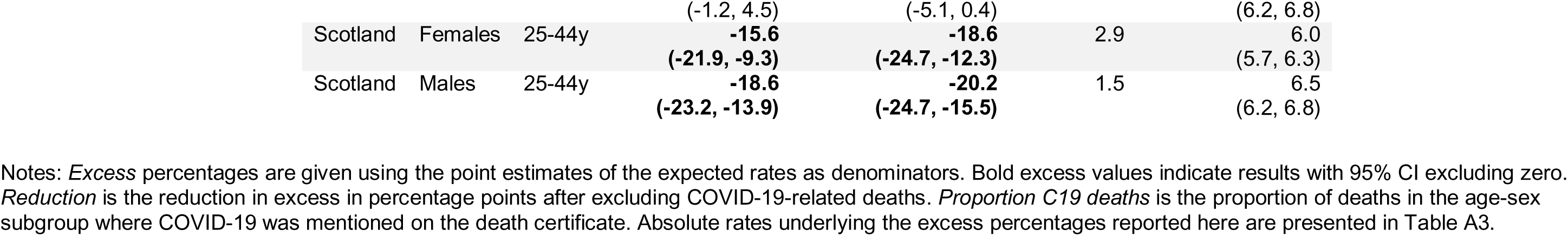
Relative excesses in observed age-standardised mortality rates per 100,000 population per year before and after excluding COVID-19-related deaths in 2022, assuming continuation of austerity-era (2012-2019) trends.

| Nation | Sex | Age Group | Excess %<br>(95% CI) | Excess % excl. C19<br>(95% CI) | Reduction | Prop. C19 deaths<br>(95% CI) |
| --- | --- | --- | --- | --- | --- | --- |
| England | Persons | All | 5.6<br>(5.3, 5.9) | -0.5<br>(-0.8, -0.2) | 6.1 | 5.7<br>(5.7, 5.8) |
| England | Females | All | 4.4<br>(4.0, 4.8) | -1.1<br>(-1.5, -0.7) | 5.5 | 5.3<br>(5.3, 5.4) |
| England | Males | All | 7.2<br>(6.8, 7.6) | 0.5<br>(0.1, 0.9) | 6.8 | 6.1<br>(6.1, 6.2) |
| England | Females | 0-74y | 7.1<br>(6.3, 7.9) | 2.5<br>(1.7, 3.3) | 4.6 | 5.3<br>(5.3, 5.4) |
| England | Males | 0-74y | 8.8<br>(8.2, 9.5) | 4.3<br>(3.6, 4.9) | 4.6 | 6.1<br>(6.1, 6.2) |
| England | Females | >74y | 6.6<br>(6.0, 7.2) | 1.1<br>(0.4, 1.7) | 5.6 | 5.3<br>(5.3, 5.4) |
| England | Males | >74y | 7.5<br>(6.9, 8.1) | 1.2<br>(0.6, 1.7) | 6.3 | 6.1<br>(6.1, 6.2) |
| England | Females | 45-64y | 6.0<br>(4.7, 7.2) | 1.8<br>(0.5, 3.0) | 4.2 | 5.3<br>(5.3, 5.4) |
| England | Males | 45-64y | 7.4<br>(6.3, 8.5) | 3.6<br>(2.6, 4.7) | 3.8 | 6.1<br>(6.1, 6.2) |
| England | Females | 25-44y | 3.2<br>(0.4, 6.0) | -0.1<br>(-2.9, 2.6) | 3.3 | 5.3<br>(5.3, 5.4) |
| England | Males | 25-44y | 4.6<br>(2.4, 6.7) | 1.9<br>(-0.2, 4.0) | 2.6 | 6.1<br>(6.1, 6.2) |
| Scotland | Persons | All | 2.9<br>(2.1, 3.7) | -3.5<br>(-4.3, -2.7) | 6.5 | 6.2<br>(6.1, 6.4) |
| Scotland | Females | All | 3.4<br>(2.2, 4.5) | -2.8<br>(-3.9, -1.6) | 6.1 | 6.0<br>(5.7, 6.3) |
| Scotland | Males | All | 2.6<br>(1.5, 3.8) | -4.3<br>(-5.4, -3.2) | 6.9 | 6.5<br>(6.2, 6.8) |
| Scotland | Females | 0-74y | 3.1<br>(1.0, 5.2) | -1.8<br>(-3.9, 0.2) | 4.9 | 6.0<br>(5.7, 6.3) |
| Scotland | Males | 0-74y | 2.2<br>(0.4, 3.9) | -2.4<br>(-4.1, -0.6) | 4.5 | 6.5<br>(6.2, 6.8) |
| Scotland | Females | >74y | 4.7<br>(2.9, 6.4) | -1.8<br>(-3.5, -0.1) | 6.5 | 6.0<br>(5.7, 6.3) |
| Scotland | Males | >74y | 4.4<br>(2.7, 6.0) | -2.3<br>(-3.9, -0.7) | 6.7 | 6.5<br>(6.2, 6.8) |
| Scotland | Females | 45-64y | 1.5<br>(-1.8, 4.9) | -2.2<br>(-5.5, 1.1) | 3.8 | 6.0<br>(5.7, 6.3) |
| Scotland | Males | 45-64y | 1.6 | -2.4 | 4.0 | 6.5 |
|  |  |  | (-1.2, 4.5) | (-5.1, 0.4) |  | (6.2, 6.8) |
| Scotland | Females | 25-44y | **-15.6** | **-18.6** | 2.9 | 6.0 |
|  |  |  | **(-21.9, -9.3)** | **(-24.7, -12.3)** |  | (5.7, 6.3) |
| Scotland | Males | 25-44y | **-18.6** | **-20.2** | 1.5 | 6.5 |
|  |  |  | **(-23.2, -13.9)** | **(-24.7, -15.5)** |  | (6.2, 6.8) |

Notes: *Excess* percentages are given using the point estimates of the expected rates as denominators. Bold excess values indicate results with 95% CI excluding zero. *Reduction* is the reduction in excess in percentage points after excluding COVID-19-related deaths. *Proportion C19 deaths* is the proportion of deaths in the age-sex subgroup where COVID-19 was mentioned on the death certificate. Absolute rates underlying the excess percentages reported here are presented in Table A3.

### Estimates of excess deaths

In England in 2022, there were 11,290 additional female and 18,450 additional male deaths compared to expected deaths based on the continuation of austerity-era trends; excluding COVID-19 deaths reduced these to −2,970 for females and 1,640 for males. In Scotland, 1,020 additional female and 790 additional male deaths occurred, falling to - 850 for females and −1,210 for males after excluding COVID-19 deaths.

Using pre-austerity trends (and after excluding COVID-19 deaths), England recorded 59,490 additional female and 82,480 additional male deaths in 2022, while Scotland recorded 4,670 additional female and 7,540 additional male deaths. Summing excess deaths from 2013–2022 yields 352,580 excess female and 451,700 excess male deaths in England (804,280 total) and 28,790 excess female and 46,360 excess male deaths in Scotland (75,150 total), amounting to 879,430 across both nations. Annual breakdowns are in Tables A4a–A4d. Excess deaths occurred across all deprivation levels (see Figures A5a and A5b).

### Sensitivity analyses

Linear regression models were fitted to 60 combinations of nation, sex, age, and area deprivation level. For each combination, three models were run, varying the start year (2011, 2012, or 2013). Expected ASMRs using 2011 or 2013 varied from −3.2% to +5.7% relative to those based on 2012. Most expected ASMRs differed by less than 1% from the 2012 results. Figure A6 in the Appendix shows the distribution of these differences.

## DISCUSSION

The findings reinforce concerns that mortality remains elevated after the peak years of the COVID-19 pandemic in the UK, even relative to changed mortality trends due to austerity. Part of the excess is due to high COVID-19 deaths in 2022, as seen elsewhere in Europe.[29–32] However, trends from 2012 to 2019 should not be accepted as normal. The change in mortality trends due to austerity was associated with an estimated 879,430 excess deaths between 2013-2022, comparable to the total number of British military deaths during World War One.[33] Although the deaths discussed in this study affected all parts of society, austerity has disproportionately harmed people in the most disadvantaged areas. Reversing austerity is crucial to mitigate the dramatic worsening of mortality trends seen since 2010.

This study’s findings broadly align with previous research employing different methods and time periods, while addressing limitations such as reliance on crude mortality or inappropriately short baseline periods that ignore important context around changing mortality trends since 2012. Pizzato and colleagues observed relative excess mortality of 5.0% in England and 5.2% in Scotland in 2022 using Poisson regression models fitted to mortality data from 2010–2019, compared to 5.6% and 2.9% in the present study.[34] Pérez-Reche used linear regression on UK crude mortality rates from 2012–2019 to predict rates for 2022 and 2023.[35] In most age groups aged 30 or over, observed excesses were significant despite uncertainty in expected rates. Although the methodologies differed, with this study using ASMRs, both studies found that the largest excesses occurred in older working age and older adults. In contrast, younger working age adults experienced more adverse trends during 2012–2019, while older groups still showed modest annual improvements.

Although international comparisons should be made cautiously due to varying pandemic progressions, higher-than-expected mortality in 2022 has been reported across Europe and worldwide.[3,29,31,32,36] Walkowiak, Domaradzki, and Walkowiak reported post-pandemic relative excess mortality in a selection of European countries up to the end of 2022.[29] They found that post-pandemic excess was negatively associated with peri-pandemic excess, ranging from around −3% in Romania to 10% in Norway, with all included Western European countries showing relative excesses of 2.5–10%. Kuhbandner and Reitzner estimated a 6.6% excess mortality in Germany for 2022, comparable to that observed in England in this study.[32] Using linear regression on 2012–2019 data, they found the 6.6% excess far exceeded predictions, even after accounting for uncertainty due to annual mortality variations. In Norway, Raknes and colleagues extrapolated all-cause ASMRs from 2010–2019 to 2022 and reported a relative excess of 13.7%, notably higher than in England and Scotland in this study.[31] The proportion of all deaths where COVID-19 was mentioned on the death certificate was, however, similar in England, Scotland, and Norway (5.7%, 6.2%, and 6.3% respectively). In line with internation results, expected ASMRs based on austerity-era trends in our study indicate that COVID-19 accounts for a substantial portion of the observed excess mortality in England and Scotland in 2022. However, any excess mortality explained by COVID-19 is dwarfed by the excess related to the change in mortality trends following the imposition of austerity which continues to accumulate even after achieving substantial control over COVID-19 mortality.

Our comparisons of recent observed ASMRs with long-term trend predictions add to and update a substantial evidence base documenting extraordinary changes in UK mortality rates since the early 2010s. These changes largely result from UK Government austerity measures implemented from 2010 onward.[16–21] UK Government spending was reduced by around £540 billion between 2010 and 2019, involving large cuts to social security and vital public services (via reductions in local government funding), which have profoundly harmed the health of the poorest and most vulnerable in society.[16] The causal pathways are well established, linking increased poverty, loss of support services, stress, poor mental health, and adverse behavioural ‘coping mechanisms’.

### Strengths and limitations

One strength of this study is its use of population-wide data for England and Scotland over two decades. Standardising ASMRs to the 2013 European Standard Population overcomes the shortcomings of raw death counts, which do not account for population changes or varying deprivation group structures. ASMRs also address the limitations of crude mortality rates that fail to adjust for changes in age and sex structure over time or between countries. By projecting ASMRs from longer pre-pandemic (2001–2010) and austerity-era (2012–2019) linear trends rather than using a five-year average, this study captures longer-term mortality shifts and provides a more accurate counterfactual than recent methods, including those published by the ONS.

This study also has limitations. ASMRs vary year-to-year, and this study did not account for that variability when defining excess mortality. Fitting linear regression models to only 11 years of data introduces uncertainty, especially when extrapolating 12 years beyond the fitting period, as it assumes a stable long-term trend that may not capture underlying fluctuations or structural changes. Using area-level deprivation measures yields smaller inequality gradients than individual measures due to substantial misclassification.[37] These indices include health outcomes which potentially introduces endogeneity, though its impact on summary measures is minimal.[27,28] Defining a direct COVID-19 death as any death mentioning COVID-19 rather than as the underlying cause may distort estimates of excess mortality from other causes. For example, in Scotland fewer than 60% of what we considered COVID-19 deaths in 2022 listed it as the underlying cause. However, many direct COVID-19 deaths are also attributable to austerity as highlighted in the ongoing public enquiry, making their exclusion from austerity-era excess mortality estimates highly conservative.[38]

### Conclusions

This study showed that mortality in 2022 remained higher than expected in England and Scotland, exceeding predictions based on both pre-pandemic austerity-era and pre-austerity trends. Our work overcomes key methodological limitations of earlier research and reveals that austerity has disproportionately impacted the most socioeconomically disadvantaged groups. The findings suggest that 879,430 excess deaths occurred between 2013 and 2022 in these nations due to changes in mortality trends linked to austerity. Although some excess mortality in 2022 is attributable to direct COVID-19 deaths, the dramatic ASMR changes since the early 2010s account for far more excess deaths yet these changes receive far less attention. Numerous studies have linked these changes to austerity. Improving UK mortality rates requires reversing the austerity-driven policies that underpin these trends, especially for those experiencing the most severe socioeconomic disadvantages.

## Supporting information

RECORD checklist

## ACKNOWLEDGMENTS

We thank National Records of Scotland for providing the mortality data and the Public Health Scotland Mortality Special Interest Group for their valuable feedback on the analysis.

## DATA AVAILABILITY STATEMENT

Data are available upon request from the Office of National Statistics and National Records of Scotland.

## CREDIT TAXONOMY

Conceptualization - All

Data curation – DB, DW

Formal Analysis – DRRB

Funding acquisition – RD, DW

Investigation - All

Methodology - All

Project administration - NA

Resources - NA

Software - NA

Supervision – DB, DW

Validation - NA

Visualization - DRRB

Writing – original draft - DRRB

Writing – review & editing – All

## FUNDING STATEMENT

This work was funded by the Glasgow Centre for Population Health. RD and DB are supported by the Medical Research Council (MC_UU_00022/2) and the Scottish Government Chief Scientist Office (SPHSU17).

The funders had no role in the design of the study or in the collection, analysis, and interpretation of data and results.

## RIGHTS RETENTION STATEMENT

For the purpose of open access, the authors have applied a Creative Commons Attribution (CC BY) licence to any Author Accepted Manuscript version arising from this submission.

The Corresponding Author has the right to grant on behalf of all authors and does grant on behalf of all authors, an exclusive licence on a worldwide basis to the BMJ Publishing Group Ltd to permit this article (if accepted) to be published in BMJ editions and any other BMJPGL products and sublicences such use and exploit all subsidiary rights, as set out in our licence.

All authors have completed the Unified Competing Interest form and declare: no support from any organisation for the submitted work; no financial relationships with any organisations that might have an interest in the submitted work in the previous three years, no other relationships or activities that could appear to have influenced the submitted work.

The lead author affirms that the manuscript is an honest, accurate, and transparent account of the study being reported; that no important aspects of the study have been omitted; and that any discrepancies from the study as planned (and, if relevant, registered) have been explained.

## Notes

### Competing Interest Statement

The authors have declared no competing interest.

### Author Declarations

Ethical approval for this work was not required.

### Summary of Updates

Minor changes to methods and introduction following peer review feedback.

