## Supplement for "Excess mortality in England and Scotland in 2022: The long shadow of austerity and the return to an unacceptable pre-pandemic baseline"

Appendix

Index of Multiple Deprivation versions

**Table A1a** – Alignment of mortality data years with the corresponding versions of the England (and Wales) Index of Multiple Deprivation (IMD)

| **Mortality data** | **IMD version** |
| --- | --- |
| 2001-05 | 2004 |
| 2006-08 | 2007 |
| 2009-12 | 2010 |
| 2013-17 | 2015 |
| 2018-22 | 2019 |

**Table A1b** – Alignment of mortality data years with the corresponding versions of the Scottish Index of Multiple Deprivation (SIMD)

| **Mortality data** | **SIMD version** |
| --- | --- |
| 2001-04 | 2004 |
| 2005-07 | 2006 |
| 2008-10 | 2009 |
| 2011-13 | 2012 |
| 2014-17 | 2016 |
| 2018-22 | 2020v2 |

Excess mortality by age subgroup

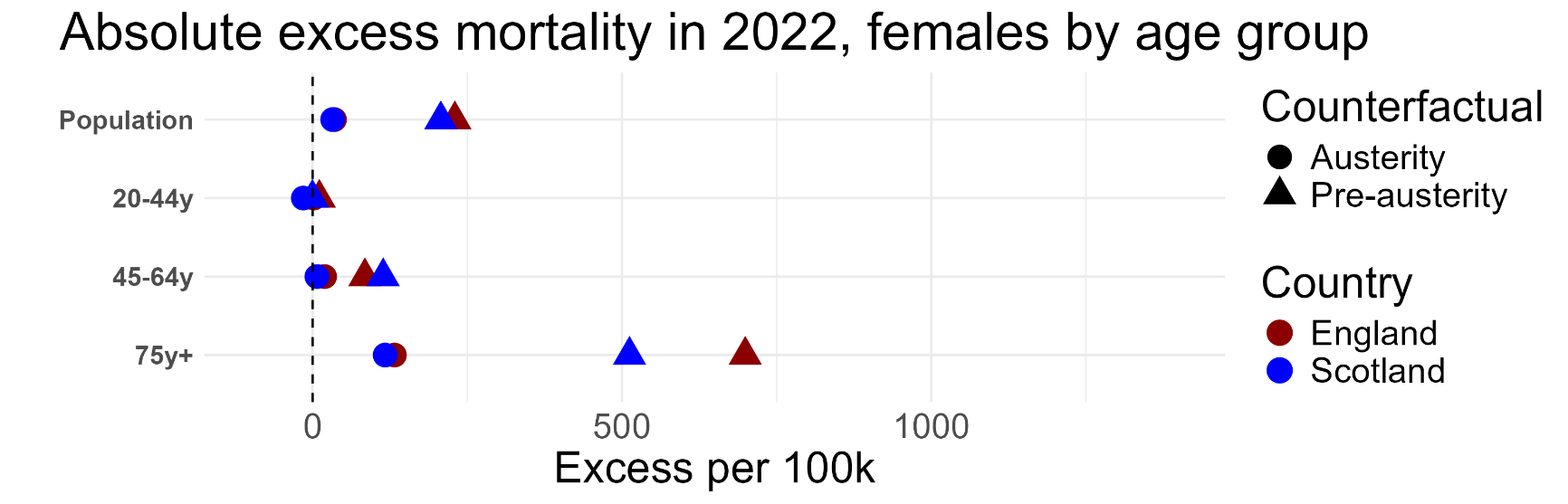

**Figure A1a** – Excess mortality across age subgroups in females based on expected ASMRs predicted by trends from austerity (2012-2019) and pre-austerity (2001-2010) eras.

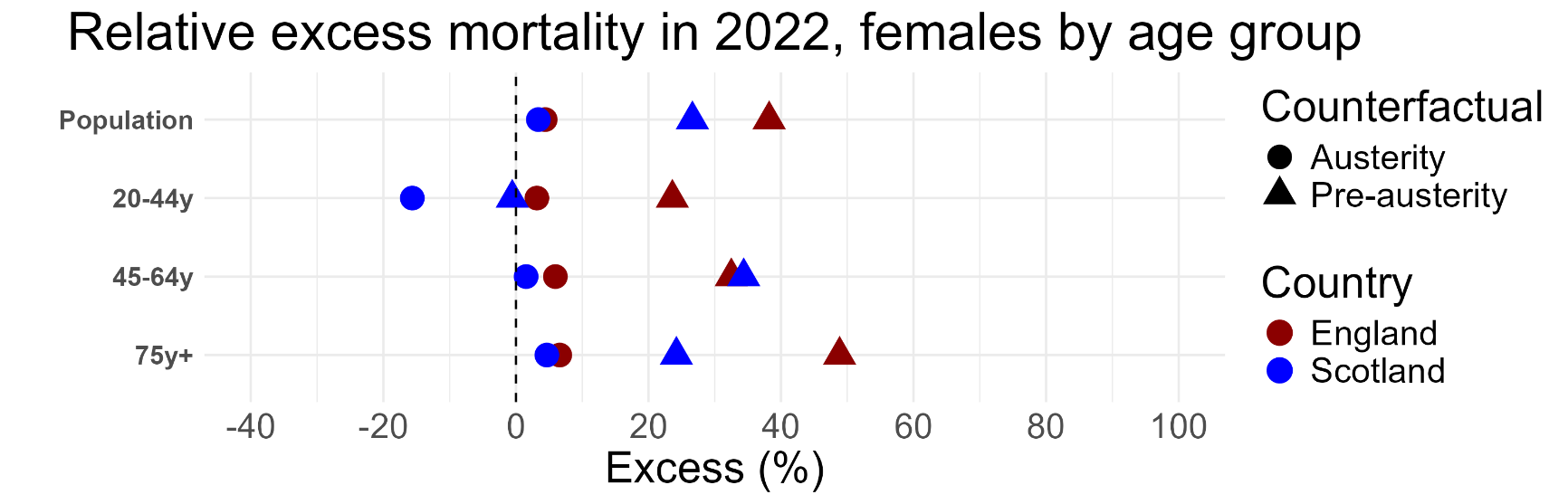

**Figure A1b** – Relative excess mortality across age subgroups in females based on expected ASMRs predicted by trends from austerity (2012-2019) and pre-austerity (2001-2010) eras.

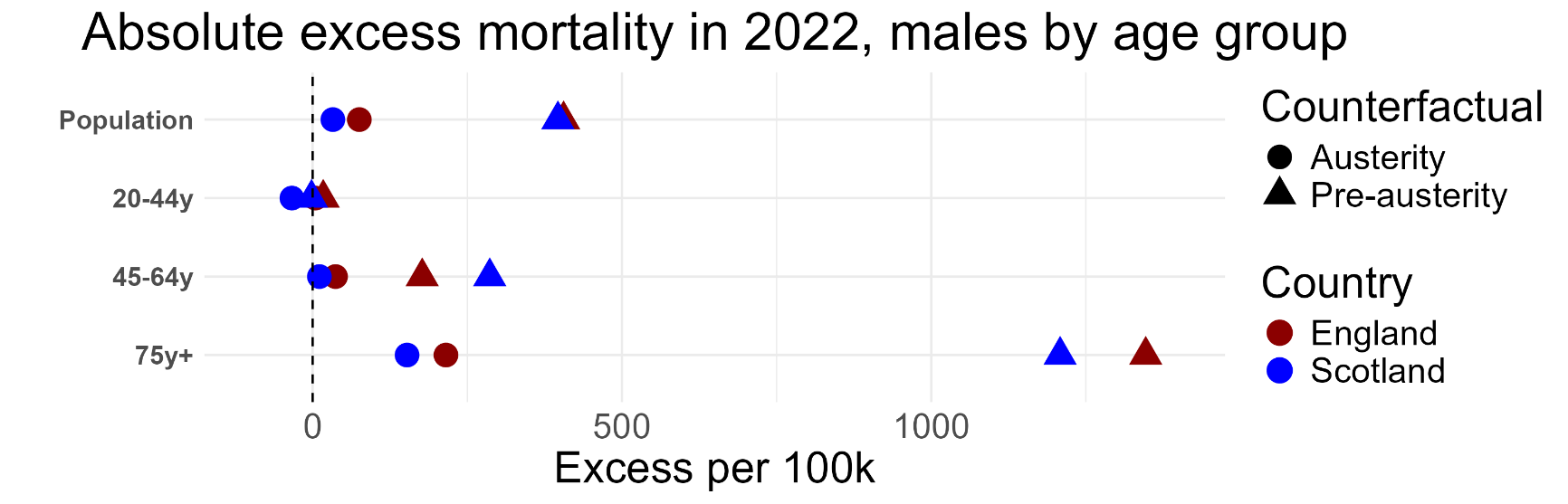

**Figure A1c** – Excess mortality across age subgroups in males based on expected ASMRs predicted by trends from austerity (2012-2019) and pre-austerity (2001-2010) eras.

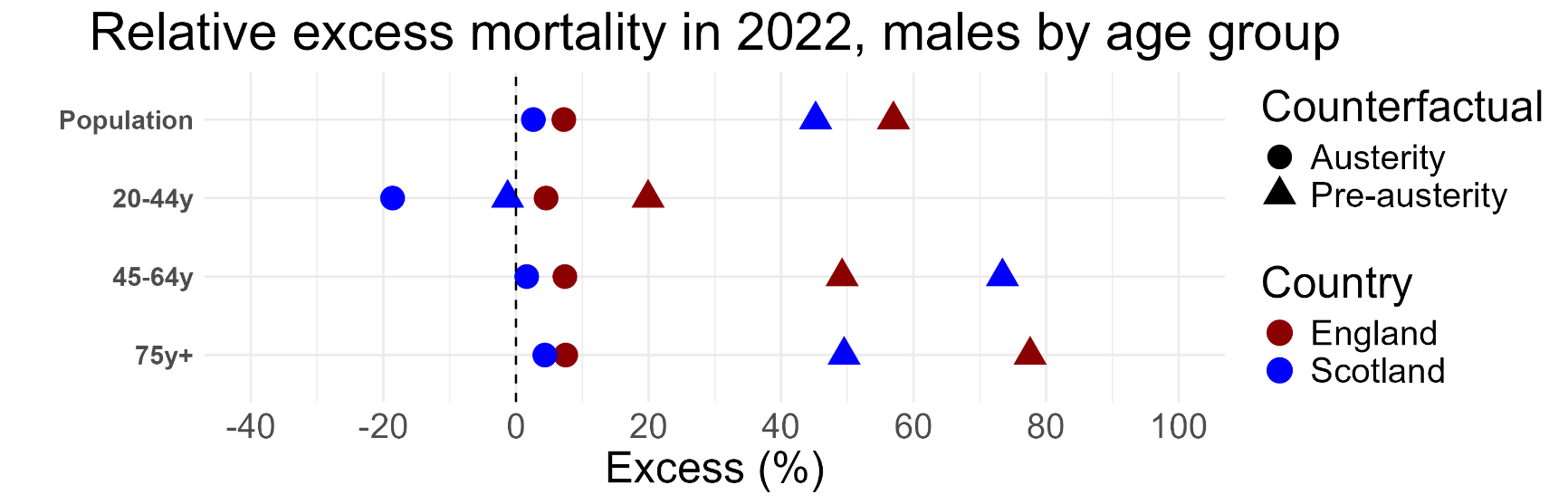

**Figure A1d** – Relative excess mortality across age subgroups in males based on expected ASMRs predicted by trends from austerity (2012-2019) and pre-austerity (2001-2010) eras.

Mortality by deprivation level

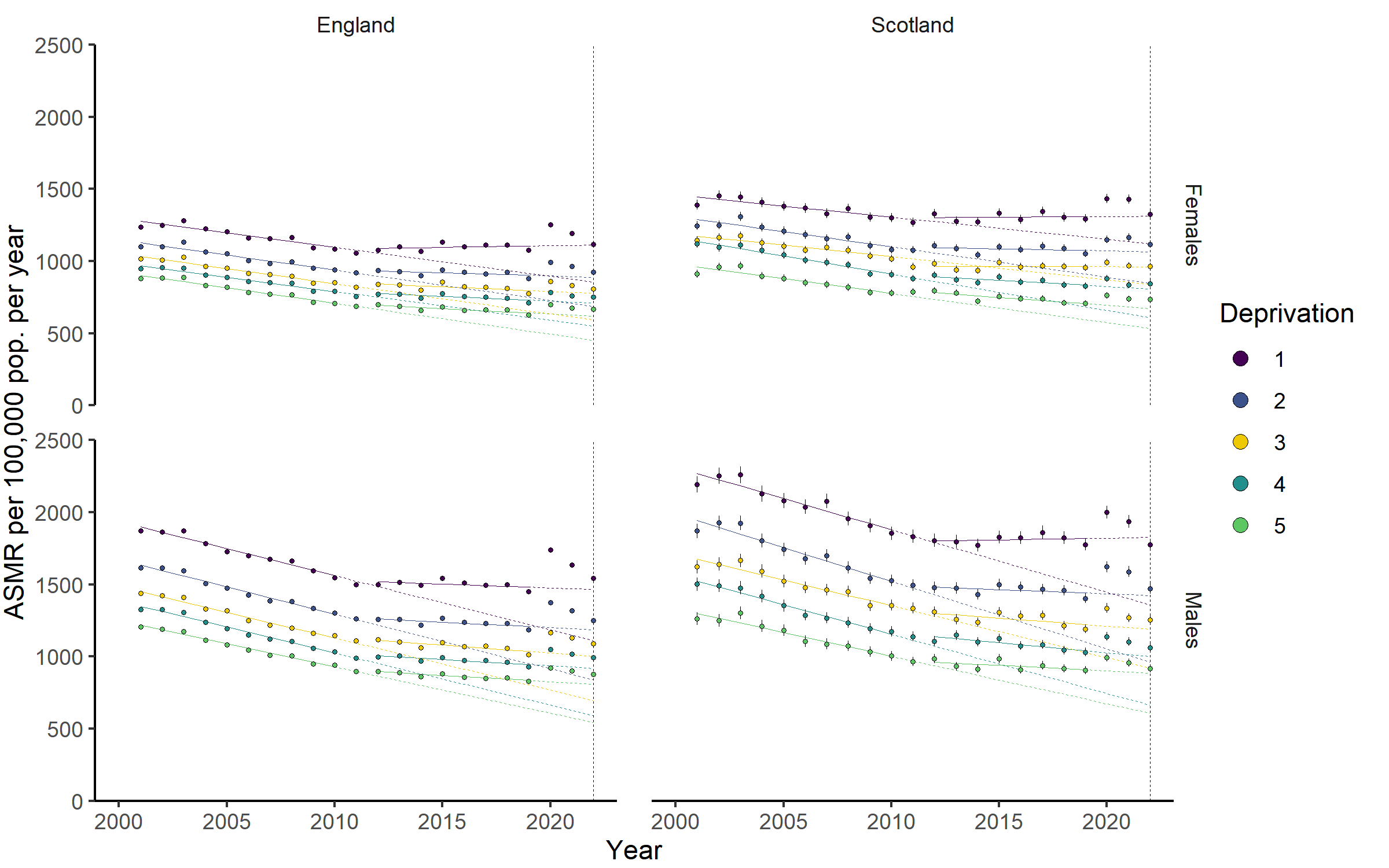

**Figure A2** - Age-standardised mortality rates from 2001–2022 by deprivation level. 1 = most deprived, 5 = least deprived. Points are observed rates. Solid lines are trends fitted through 2001–2010 (pre-austerity) and 2012–2019 (austerity imposed). Dashed lines are trends projected to 2022. Confidence intervals are included for observed rates but are so narrow as to be obscured by the points in most cases.

**Table A2a** – Age-standardised all-cause mortality rates, rate ratios, absolute excess in ASMRs, and relative excess in ASMRs in 2022 by fifths of deprivation, assuming continuation of austerity era (2012-2019) trends.

| **Nation** | **Sex** | **Deprivation** | **Obs. Rate (95% CI)** | **Rate Ratio** | **Exp. Rate** | **Excess % (95% CI)** |
| --- | --- | --- | --- | --- | --- | --- |
| England | Females | 1 | 1117 (1108, 1127) | 1.68 | 1112 | 0.5 (-0.4, 1.4) |
| England | Females | 2 | 924 (916, 932) | 1.39 | 889 | 4.0 (3.1, 4.8) |
| England | Females | 3 | 808 (801, 815) | 1.21 | 778 | 3.9 (3.1, 4.8) |
| England | Females | 4 | 749 (743, 756) | 1.12 | 710 | 5.6 (4.7, 6.4) |
| England | Females | 5 | 667 (661, 673) |  | 618 | 7.9 (6.9, 8.8) |
| England | Males | 1 | 1541 (1528, 1554) | 1.75 | 1465 | 5.2 (4.3, 6.1) |
| England | Males | 2 | 1249 (1238, 1259) | 1.42 | 1184 | 5.4 (4.5, 6.3) |
| England | Males | 3 | 1088 (1079, 1097) | 1.24 | 1002 | 8.6 (7.7, 9.5) |
| England | Males | 4 | 992 (984, 1000) | 1.13 | 919 | 8.0 (7.1, 8.9) |
| England | Males | 5 | 878 (871, 886) |  | 809 | 8.6 (7.7, 9.5) |
| Scotland | Females | 1 | 1324 (1294, 1356) | 1.80 | 1311 | 1.0 (-1.4, 3.4) |
| Scotland | Females | 2 | 1118 (1091, 1144) | 1.52 | 1066 | 4.8 (2.4, 7.3) |
| Scotland | Females | 3 | 965 (942, 989) | 1.31 | 962 | 0.4 (-2.0, 2.9) |
| Scotland | Females | 4 | 845 (823, 868) | 1.15 | 809 | 4.5 (1.8, 7.3) |
| Scotland | Females | 5 | 735 (715, 756) |  | 673 | 9.3 (6.3, 12.4) |
| Scotland | Males | 1 | 1774 (1733, 1816) | 1.94 | 1826 | -2.8 (-5.1, -0.5) |
| Scotland | Males | 2 | 1470 (1435, 1506) | 1.61 | 1423 | 3.4 (0.9, 5.9) |
| Scotland | Males | 3 | 1255 (1224, 1286) | 1.37 | 1189 | 5.6 (3.0, 8.2) |
| Scotland | Males | 4 | 1062 (1034, 1091) | 1.16 | 999 | 6.3 (3.5, 9.2) |
| Scotland | Males | 5 | 915 (888, 942) |  | 886 | 3.2 (0.2, 6.3) |

Notes: *Depr.* 1 = most deprived areas, 5 = least deprived areas. Rates are per 100,000 population. *Rate ratio* is the ratio of observed ASMR compared to the appropriate least deprived fifth of areas. *Exp. rate* is the expected ASMRs if linear trends from the period 2012-2019 had continued to 2022 (exemplified by dashed lines in Figures 1, 2, and A2). *Excess* percentages are given using the expected rates as denominators. Bold excess values indicate results where the 95% CI of the relative excess excludes zero.

**Table A2b** – Age-standardised all-cause mortality rates, rate ratios, absolute excess in ASMRs, and relative excess in ASMRs in 2022 by fifths of deprivation, assuming continuation of austerity era (2001-2010) trends.

| **Nation** | **Sex** | **Deprivation** | **Obs. Rate (95% CI)** | **Rate Ratio** | **Exp. Rate** | **Excess % (95% CI)** |
| --- | --- | --- | --- | --- | --- | --- |
| England | Females | 1 | 1117 (1108, 1127) | 1.68 | 855 | 30.7 (29.6, 31.8) |
| England | Females | 2 | 924 (916, 932) | 1.39 | 687 | 34.6 (33.4, 35.7) |
| England | Females | 3 | 808 (801, 815) | 1.21 | 593 | 36.2 (35.1, 37.4) |
| England | Females | 4 | 749 (743, 756) | 1.12 | 551 | 36.1 (35.0, 37.3) |
| England | Females | 5 | 667 (661, 673) |  | 452 | 47.5 (46.2, 48.8) |
| England | Males | 1 | 1541 (1528, 1554) | 1.75 | 1110 | 38.8 (37.7, 40.0) |
| England | Males | 2 | 1249 (1238, 1259) | 1.42 | 839 | 48.7 (47.5, 50.0) |
| England | Males | 3 | 1088 (1079, 1097) | 1.24 | 697 | 56.0 (54.7, 57.3) |
| England | Males | 4 | 992 (984, 1000) | 1.13 | 593 | 67.4 (66.0, 68.8) |
| England | Males | 5 | 878 (871, 886) |  | 543 | 61.8 (60.4, 63.2) |
| Scotland | Females | 1 | 1324 (1294, 1356) | 1.80 | 1122 | 18.1 (15.3, 20.8) |
| Scotland | Females | 2 | 1118 (1091, 1144) | 1.52 | 847 | 32.0 (28.9, 35.2) |
| Scotland | Females | 3 | 965 (942, 989) | 1.31 | 838 | 15.2 (12.4, 18.0) |
| Scotland | Females | 4 | 845 (823, 868) | 1.15 | 609 | 38.9 (35.2, 42.5) |
| Scotland | Females | 5 | 735 (715, 756) |  | 533 | 37.9 (34.1, 41.8) |
| Scotland | Males | 1 | 1774 (1733, 1816) | 1.94 | 1359 | 30.6 (27.5, 33.7) |
| Scotland | Males | 2 | 1470 (1435, 1506) | 1.61 | 961 | 53.0 (49.4, 56.8) |
| Scotland | Males | 3 | 1255 (1224, 1286) | 1.37 | 922 | 36.1 (32.8, 39.5) |
| Scotland | Males | 4 | 1062 (1034, 1091) | 1.16 | 663 | 60.3 (56.0, 64.7) |
| Scotland | Males | 5 | 915 (888, 942) |  | 608 | 50.5 (46.1, 54.9) |

Excluding COVID-19-related deaths

**Table A3** - Age-standardised mortality rates before and after excluding COVID-19-related deaths in 2022, supporting Table 3.

| **Nation** | **Sex** | **Age group** | **Exp. rate**  **(95% CI)** | **Obs. rate**  **(95% CI)** | **Obs. rate exc. C19**  **(95% CI)** |
| --- | --- | --- | --- | --- | --- |
| England | Persons | All | 911 | 961 (959, 964) | **906 (904, 909)** |
| England | Females | All | 796 | 831 (828, 835) | **787 (784, 791)** |
| England | Males | All | 1042 | 1118 (1114, 1122) | **1047 (1043, 1051)** |
| England | Females | 0-74y | 251 | 269 (267, 271) | **258 (256, 260)** |
| England | Males | 0-74y | 386 | 420 (417, 422) | **402 (400, 405)** |
| England | Females | >74y | 1999 | 2131 (2118, 2144) | **2020 (2008, 2032)** |
| England | Males | >74y | 2867 | 3082 (3066, 3099) | **2901 (2885, 2917)** |
| England | Females | 45-64y | 325 | 344 (340, 348) | **330 (326, 334)** |
| England | Males | 45-64y | 500 | 537 (532, 542) | **518 (513, 523)** |
| England | Females | 24-44y | 55 | 56 (55, 58) | **55 (53, 56)** |
| England | Males | 24-44y | 98 | 103 (100, 105) | **100 (98, 102)** |
| Scotland | Persons | All | 1084 | 1116 (1107, 1125) | **1046 (1038, 1055)** |
| Scotland | Females | All | 953 | 985 (974, 996) | **926 (916, 937)** |
| Scotland | Males | All | 1240 | 1273 (1259, 1288) | **1187 (1173, 1201)** |
| Scotland | Females | 0-74y | 337 | 347 (340, 354) | **331 (324, 337)** |
| Scotland | Males | 0-74y | 503 | 514 (505, 523) | **491 (483, 500)** |
| Scotland | Females | >74y | 2510 | 2628 (2584, 2672) | **2465 (2423, 2508)** |
| Scotland | Males | >74y | 3496 | 3648 (3592, 3705) | **3415 (3360, 3470)** |
| Scotland | Females | 45-64y | 439 | 446 (431, 460) | **429 (415, 444)** |
| Scotland | Males | 45-64y | 665 | 676 (658, 695) | **650 (631, 668)** |
| Scotland | Females | 24-44y | 97 | 82 (76, 88) | 79 (73, 85) |
| Scotland | Males | 24-44y | 181 | 147 (139, 155) | 144 (136, 152) |

Notes: *Exp. rate* is the expected ASMR per 100,000 population assuming the continuation of linear trends from the period 2012-2019 to 2022 (exemplified by dashed lines in Figures 1, 2, and A2). *Obs. rate* is the observed ASMR per 100,000 population and *Obs. rate exc. C19* is the ASMR per 100,000 population where deaths were removed from the numerator if COVID-19 was mentioned on the death certificate. Bold figures in the *Obs. rate exc. C19* column indicate this rate is significantly lower than the observed all-cause ASMR (*Obs. rate*).

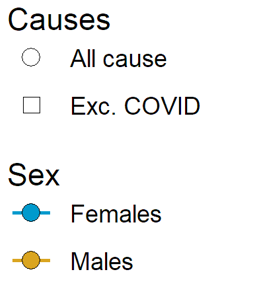

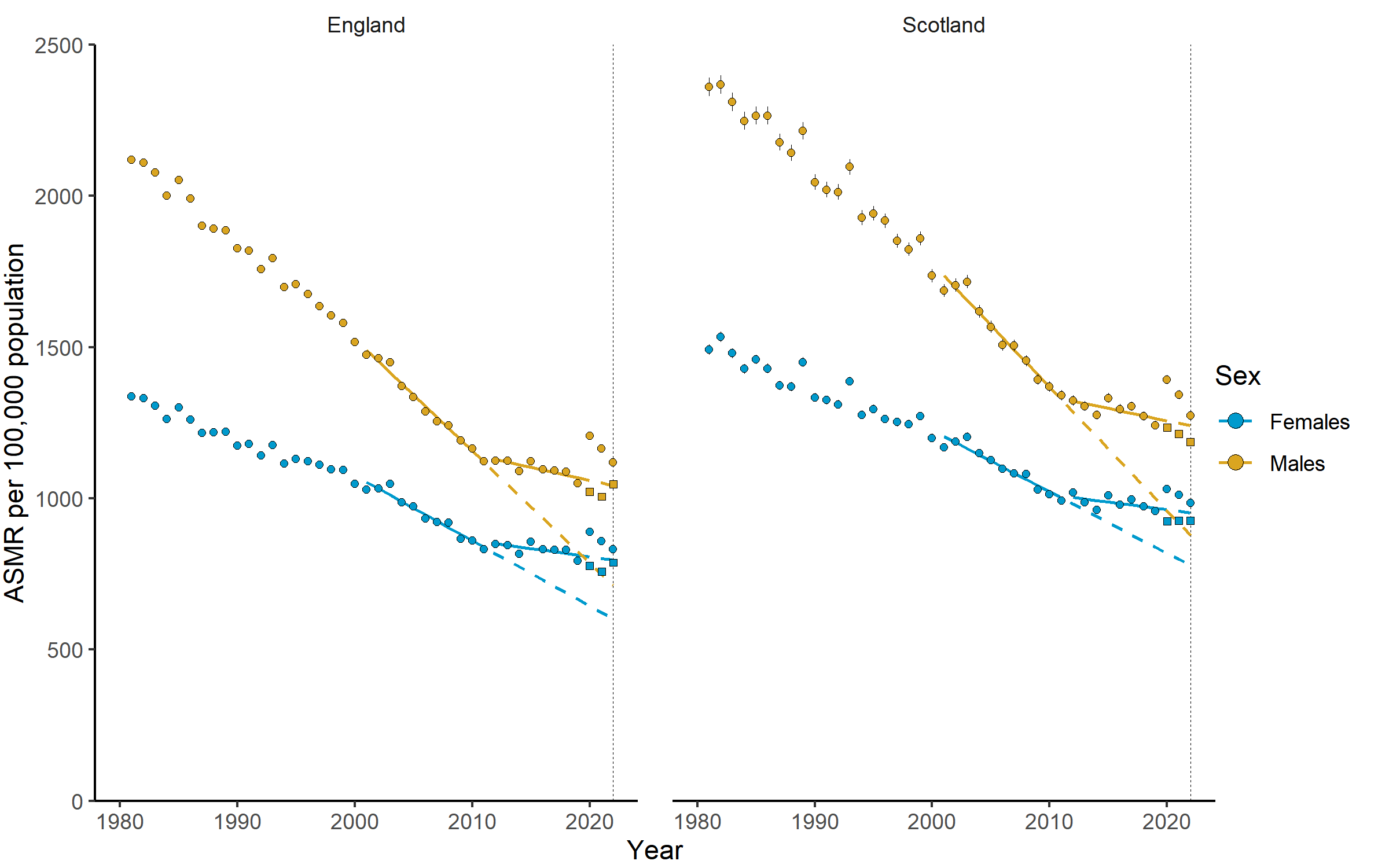

**Figure A3** - Age-standardised mortality rates from 2010–2022 for all-cause mortality and all-cause excluding cases where COVID-19 was mentioned on the death certificate. Points are observed rates. Square points are observed rates excluding COVID-19 deaths. Solid lines are trends fitted through 2001–2010 (pre-austerity). Dashed lines are trends projected to 2022. Confidence intervals are included for observed rates but are so narrow as to be obscured by the points in most cases.

Estimating absolute excess death counts

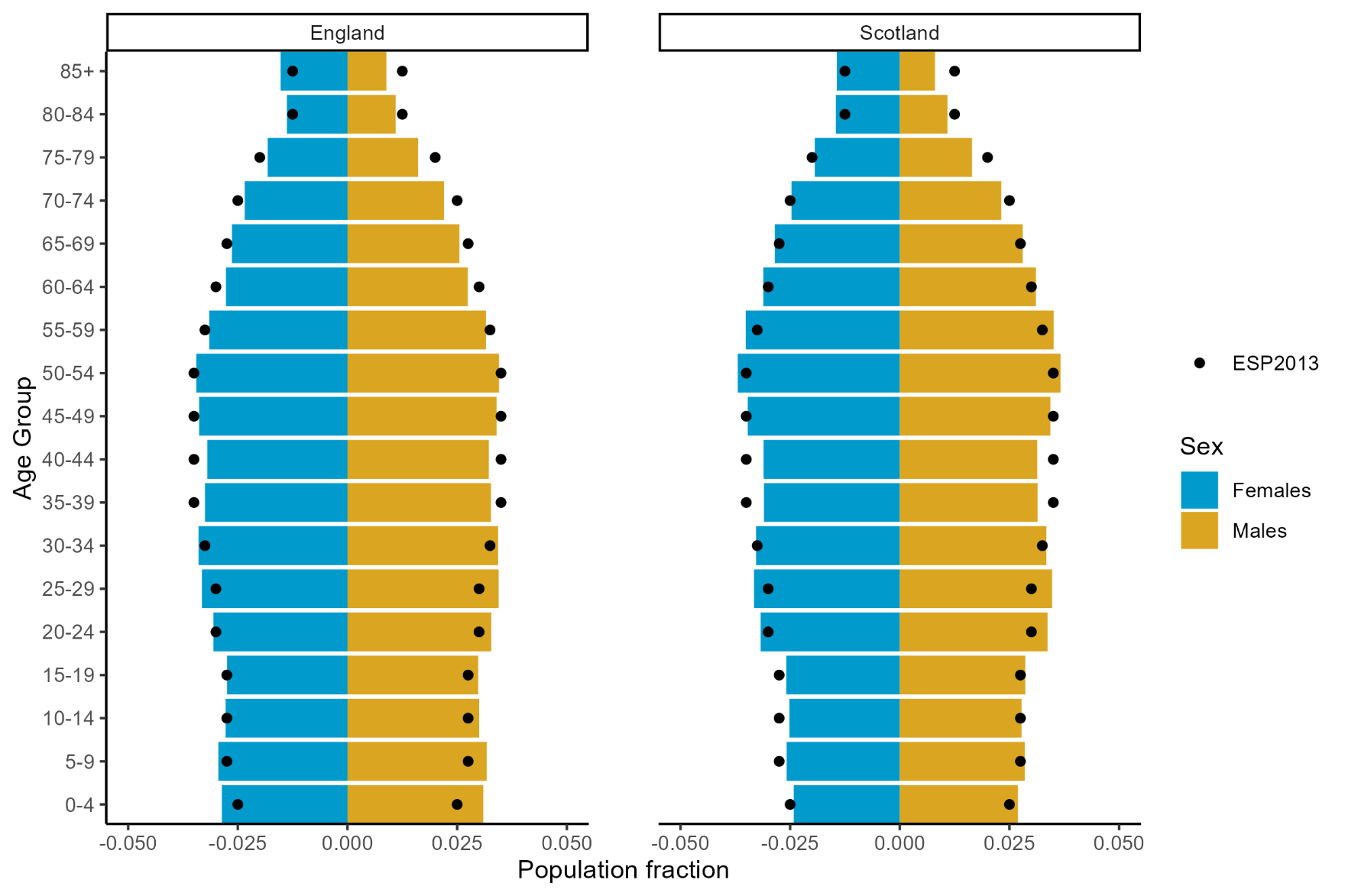

**Figure A4** – Mean fractional population distribution by five-year age group for 2013-2022 (inclusive) in England and Scotland compared to the European Standard Population 2013. Shows face validity of multiplying national population by mortality rates standardised to the European Standard Population to estimate absolute deaths.

Numerical example of excess deaths counts – Females in England in 2020

- Observed deaths = 280,958 (presented in column *Deaths* in the tables below)
- Observed ESP2013-standardised rate * population = (888.357 /100,000 * 28,567,320) = 253,779.8 (presented in column *Obs. rate * pop*)
- ESP2013 correction factor = 280,958 / 253,779.8 = 1.107094 (presented in column *Ratio*)
- Expected deaths in 2020 based on 2001-2010 trend continuing = Expected rate * population * ESP2013 correction factor
  = 644.6 * 28,567,320 * 1.107094 = 203,858 (presented in column *Exp. deaths (scaled)*)
- Excess deaths = observed deaths – COVID19 deaths – expected deaths
  = 280,958 – 35,483 – 203,857.7 = 41,618 (allowing for rounding in the ESP2013 correction factor and expected deaths; presented in column *Excess deaths*)

**Table A4a** - Estimates of absolute excess female deaths in England in 2013-2022 had 2001-2010 trends had continued.

| **Year** | **Deaths** | **Obs. rate * pop.** | **Ratio** | **C19 deaths** | **Deaths exc. C19** | **Exp. deaths (unscaled)** | **Exp. deaths (scaled)** | **Excess deaths** |
| --- | --- | --- | --- | --- | --- | --- | --- | --- |
| 2013 | 244261 | 231129 | 1.06 | 0 | 244261 | 217360 | 229710 | 14551 |
| 2014 | 239759 | 224933 | 1.07 | 0 | 239759 | 213113 | 227160 | 12599 |
| 2015 | 254892 | 237901 | 1.07 | 0 | 254892 | 208791 | 223703 | 31189 |
| 2016 | 250070 | 232651 | 1.07 | 0 | 250070 | 204351 | 219651 | 30419 |
| 2017 | 253418 | 233629 | 1.08 | 0 | 253418 | 199545 | 216447 | 36971 |
| 2018 | 255847 | 234846 | 1.09 | 0 | 255847 | 194662 | 212070 | 43777 |
| 2019 | 248476 | 225493 | 1.1 | 0 | 248476 | 189567 | 208888 | 39588 |
| 2020 | 280958 | 253780 | 1.11 | 35483 | 245475 | 184138 | 203858 | 41618 |
| 2021 | 270176 | 247738 | 1.09 | 31736 | 238440 | 179775 | 196057 | 42383 |
| 2022 | 266754 | 242120 | 1.1 | 14260 | 252494 | 175182 | 193005 | 59489 |
| Totals | 2564611 | - | - | 81479 | - | 1966484 | 2130549 | 352584 |

*Deaths* = observed deaths, *Obs. rate * pop.* = observed ESP2013-standardised * population, *Ratio* = correction factor used to adjust for differences in age distribution between population under observation and ESP2013 notional population, *C19 deaths* = observed deaths where COVID-19 was mentioned on the death certificate, *Deaths exc. C19* = observed deaths excluding deaths where COVID-19 was noted on the death certificate, *Exp. deaths (unscaled)* = expected ESP2013-standardised rate multiplied by population, *Exp. deaths (scaled)* = expected ESP2013-standardised rate multiplied by population multiplied by ESP2013 correction factor (Ratio), *Excess deaths* = observed deaths – C19 deaths – Exp. deaths (scale)

**Table A4b** – Estimates of absolute excess male deaths in England in 2013-2022 had 2001-2010 trends had continued.

| **Year** | **Deaths** | **Obs. * pop.** | **Ratio** | **C19 deaths** | **Deaths exc. C19** | **Exp. deaths (unscaled)** | **Exp. deaths (scaled)** | **Excess deaths** |
| --- | --- | --- | --- | --- | --- | --- | --- | --- |
| 2013 | 229291 | 298127 | 0.77 | 0 | 229291 | 277399 | 213349 | 15942 |
| 2014 | 229118 | 291933 | 0.78 | 0 | 229118 | 269983 | 211891 | 17227 |
| 2015 | 240417 | 303484 | 0.79 | 0 | 240417 | 262554 | 207992 | 32425 |
| 2016 | 240721 | 299171 | 0.8 | 0 | 240721 | 255080 | 205244 | 35477 |
| 2017 | 245465 | 299987 | 0.82 | 0 | 245465 | 246584 | 201768 | 43697 |
| 2018 | 250012 | 300954 | 0.83 | 0 | 250012 | 238013 | 197725 | 52287 |
| 2019 | 247894 | 292306 | 0.85 | 0 | 247894 | 229081 | 194275 | 53619 |
| 2020 | 288742 | 337880 | 0.85 | 43793 | 244949 | 219992 | 187998 | 56950 |
| 2021 | 279173 | 322263 | 0.87 | 37926 | 241247 | 207376 | 179648 | 61599 |
| 2022 | 273579 | 312776 | 0.87 | 16808 | 256771 | 199265 | 174293 | 82477 |
| - | 2524412 | - | - | 98527 | - | 2405327 | 1974183 | 451700 |

**Table A4c** - Estimates of absolute excess female deaths in Scotland in 2013-2022 had 2001-2010 trends had continued.

| **Year** | **Deaths** | **Obs. * pop.** | **Ratio** | **C19 deaths** | **Deaths exc. C19** | **Exp. deaths (unscaled)** | **Exp. deaths (scaled)** | **Excess deaths** |
| --- | --- | --- | --- | --- | --- | --- | --- | --- |
| 2013 | 28127 | 27061 | 1.04 | 0 | 28127 | 26341 | 27378 | 749 |
| 2014 | 27869 | 26437 | 1.05 | 0 | 27869 | 25876 | 27278 | 591 |
| 2015 | 29485 | 27861 | 1.06 | 0 | 29485 | 25422 | 26904 | 2581 |
| 2016 | 28938 | 27184 | 1.06 | 0 | 28938 | 24992 | 26605 | 2333 |
| 2017 | 29896 | 27755 | 1.08 | 0 | 29896 | 24491 | 26381 | 3515 |
| 2018 | 29452 | 27149 | 1.08 | 0 | 29452 | 23967 | 26000 | 3452 |
| 2019 | 29508 | 26834 | 1.1 | 0 | 29508 | 23491 | 25832 | 3676 |
| 2020 | 31947 | 28843 | 1.11 | 3344 | 28603 | 22926 | 25392 | 3211 |
| 2021 | 31719 | 28399 | 1.12 | 2686 | 29033 | 22408 | 25028 | 4005 |
| 2022 | 31085 | 27590 | 1.13 | 1863 | 29222 | 21788 | 24548 | 4674 |
| - | 298026 | - | - | 7893 | - | 241702 | 261346 | 28787 |

**Table A4d** - Estimates of absolute excess male deaths in Scotland in 2013-2022 had 2001-2010 trends had continued.

| **Year** | **Deaths** | **Obs. * pop.** | **Ratio** | **C19 deaths** | **Deaths exc. C19** | **Exp. deaths (unscaled)** | **Exp. deaths (scaled)** | **Excess deaths** |
| --- | --- | --- | --- | --- | --- | --- | --- | --- |
| 2013 | 26033 | 33732 | 0.77 | 0 | 26033 | 32202 | 24852 | 1181 |
| 2014 | 26094 | 33120 | 0.79 | 0 | 26094 | 31264 | 24632 | 1462 |
| 2015 | 27639 | 34749 | 0.8 | 0 | 27639 | 30363 | 24151 | 3488 |
| 2016 | 27659 | 34034 | 0.81 | 0 | 27659 | 29486 | 23963 | 3696 |
| 2017 | 28342 | 34448 | 0.82 | 0 | 28342 | 28549 | 23488 | 4854 |
| 2018 | 28253 | 33674 | 0.84 | 0 | 28253 | 27556 | 23120 | 5133 |
| 2019 | 28259 | 33063 | 0.85 | 0 | 28259 | 26615 | 22748 | 5511 |
| 2020 | 32105 | 37088 | 0.87 | 3496 | 28609 | 25546 | 22114 | 6495 |
| 2021 | 31617 | 35879 | 0.88 | 3006 | 28611 | 24523 | 21610 | 7001 |
| 2022 | 30599 | 33695 | 0.91 | 1989 | 28610 | 23202 | 21070 | 7540 |
| - | 286600 | - | - | 8491 | - | 279306 | 231748 | 46361 |

Distribution of excess deaths by deprivation level

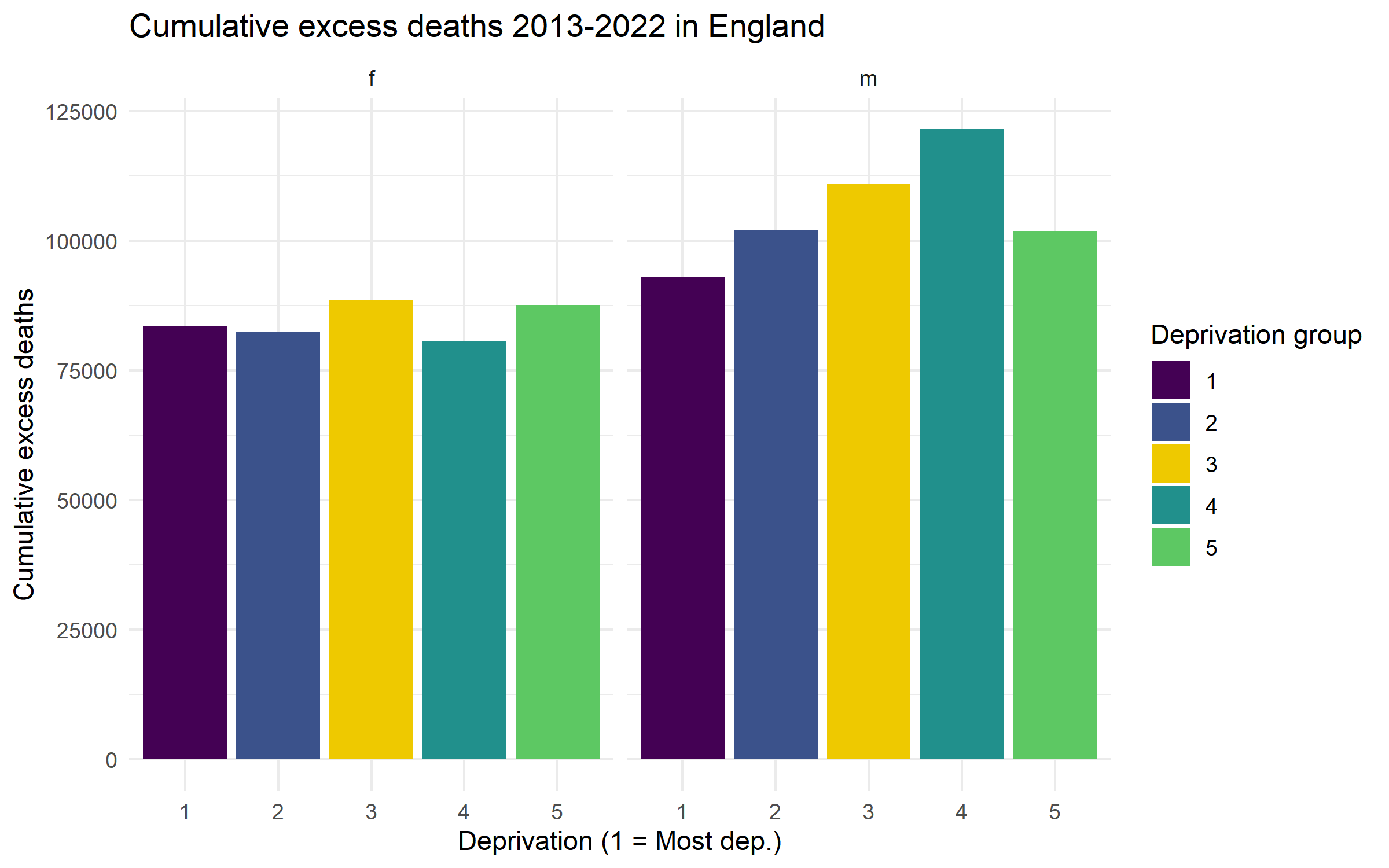

**Figure A5a** – Distribution of total estimated excess deaths by deprivation and sex in England. Cumulative totals over 2013—2022 inclusive.

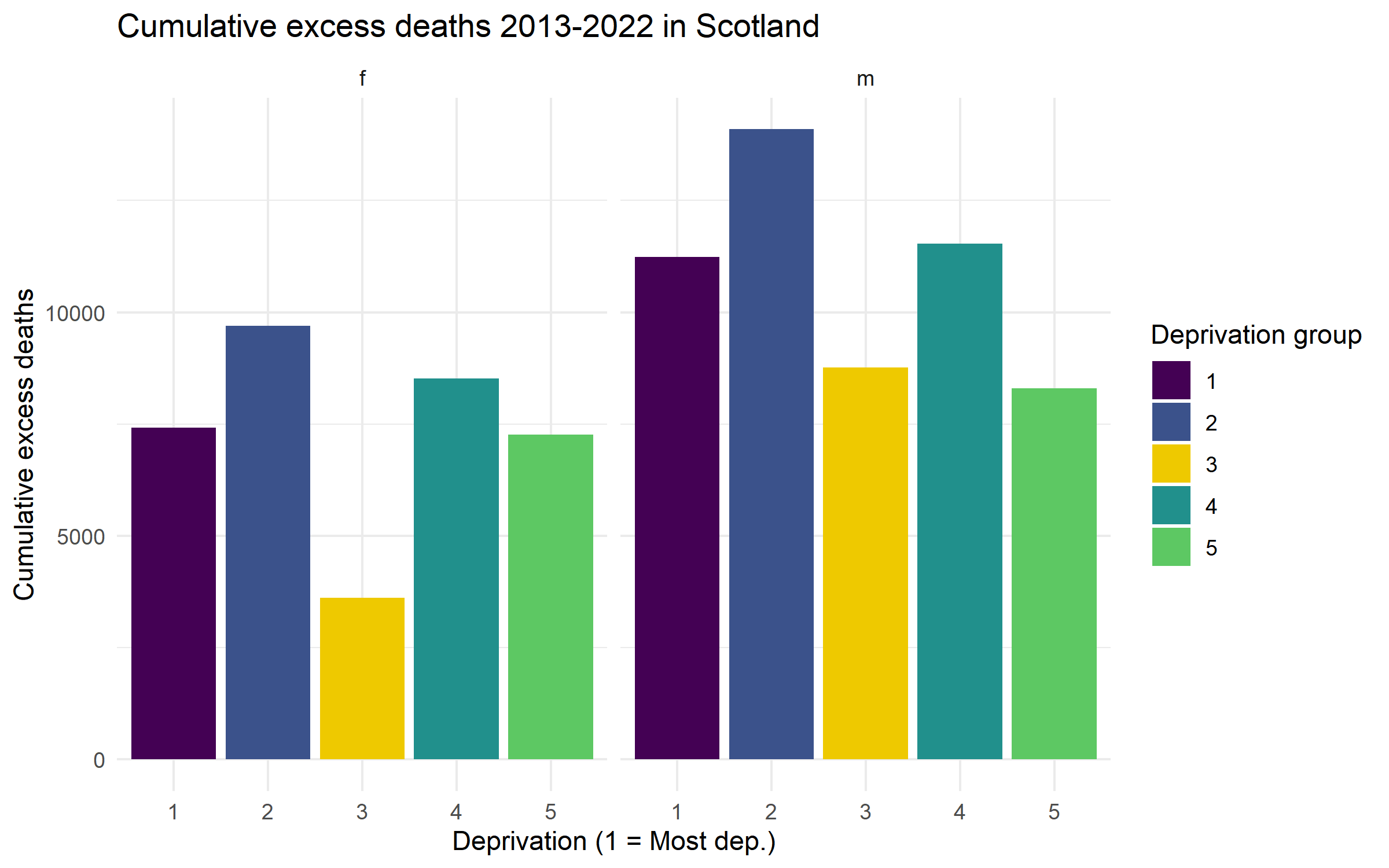

**Figure A5b** – Distribution of total estimated excess deaths by deprivation and sex in Scotland. Cumulative totals over 2013—2022 inclusive.

Sensitivity analysis – Start year

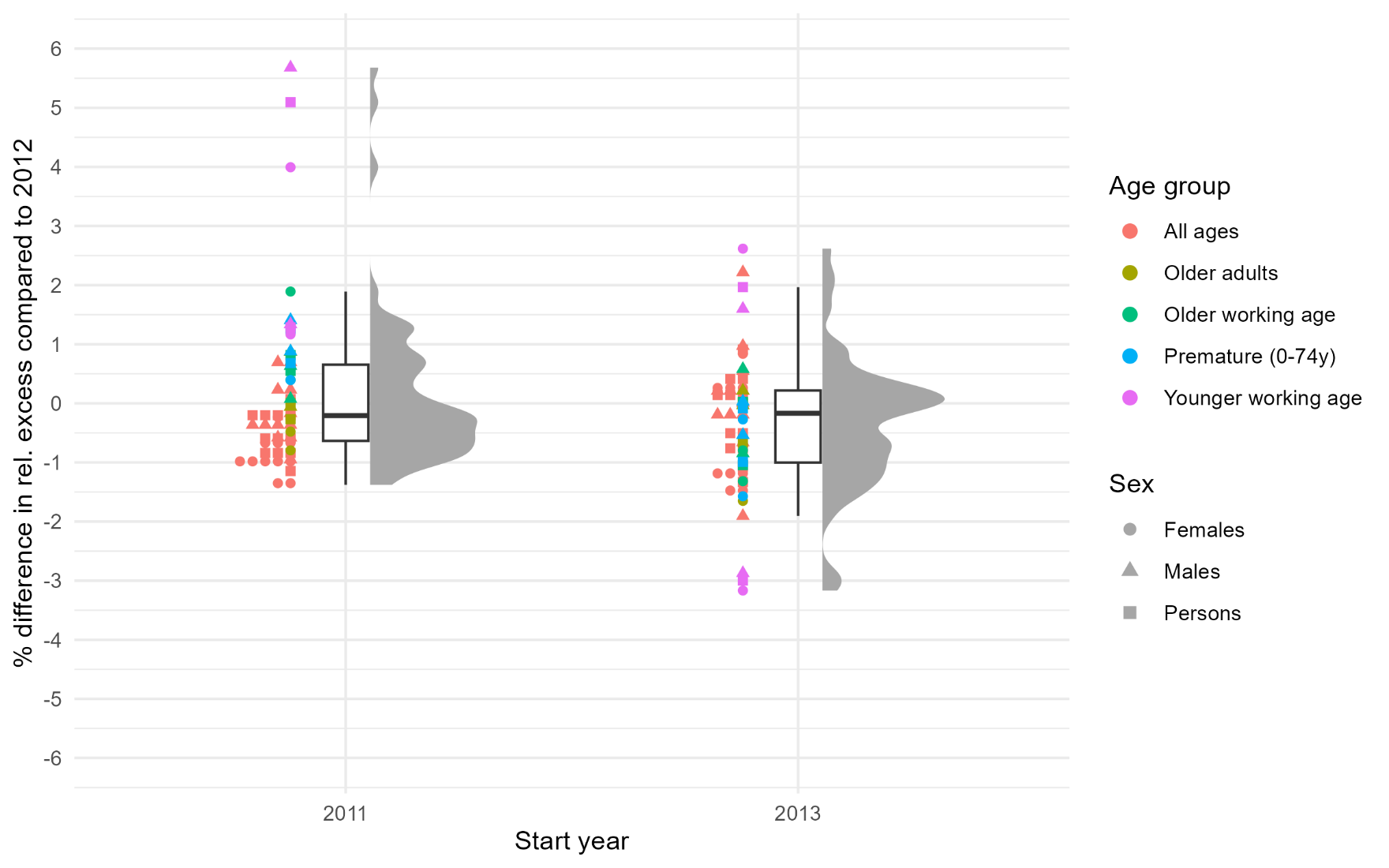

**Figure A6** – Differences in relative excess mortality values across combinations of age group and sex when using 2011 or 2013 as the start year for linear regression models compared to using 2012.

COVID-19 infections in England in 2022

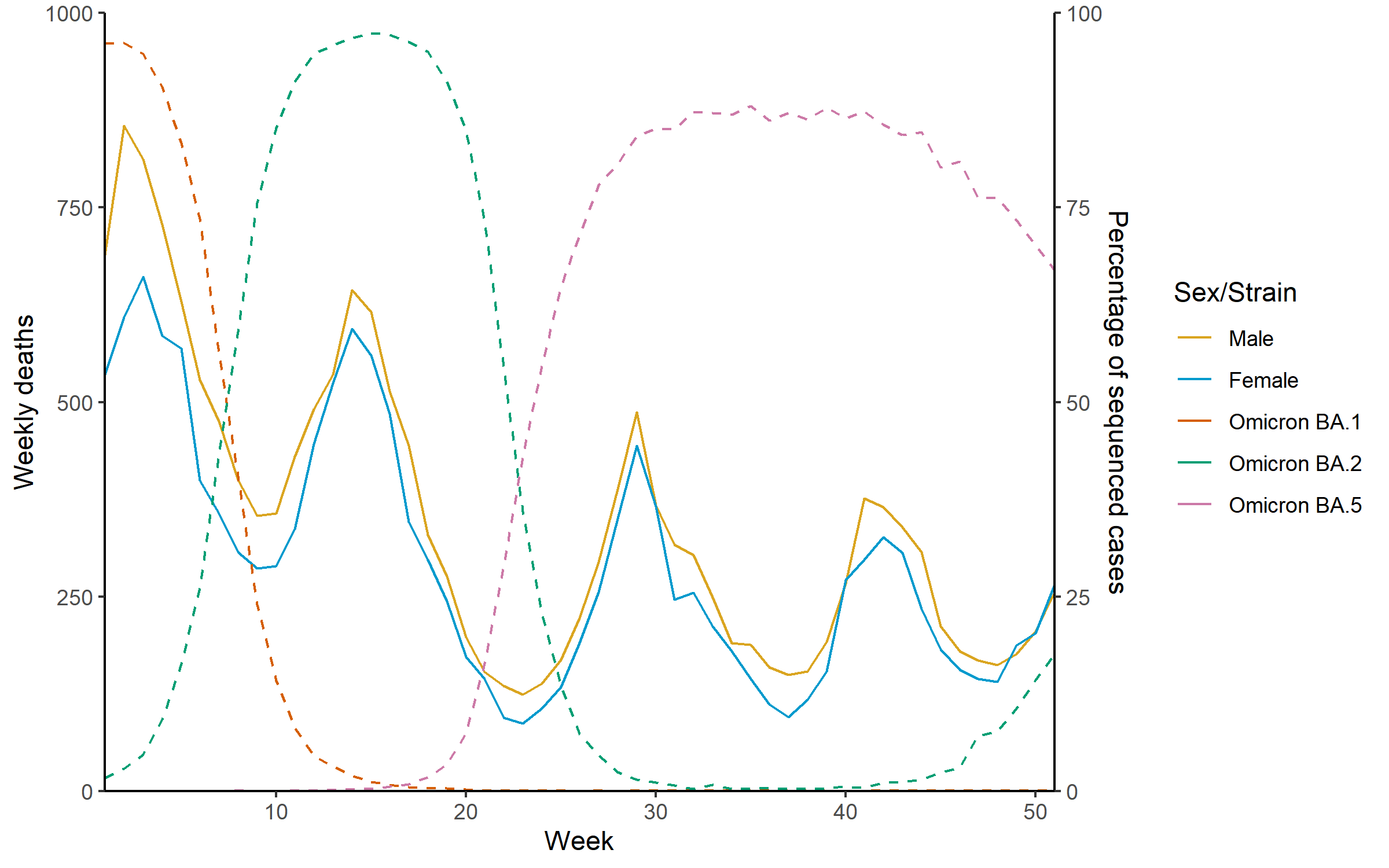

**Figure A7** – Weekly deaths in England by sex in 2022 (left vertical axis) and proportion of sequenced COVID-19 samples by strain (right vertical axis) [39]
